# Boora, an AI-assisted digital platform for overweight and obesity care in Brazilian primary care: a formative mixed-methods evaluation of perceived usability and acceptability

**DOI:** 10.64898/2026.07.15.26358116

**Authors:** Felipe da Fonseca Silva Couto, Carlos Podalirio Borges de Almeida

**Affiliations:** Professional Master’s Program in Family Health (PROFSAÚDE), Institute of Health and Biological Studies, Federal University of the South and Southeast of Pará (UNIFESSPA), Avenida dos Ipês, s/n, Cidade Universitária, Loteamento Cidade Jardim, Marabá 68508-970, Pará, Brazil; Department of Health Management, School of Nursing, Federal University of Minas Gerais (UFMG), Av. Prof. Alfredo Balena, 190, Room 530, Santa Efigênia, Belo Horizonte, Minas Gerais 30130-100, Brazil

**Keywords:** Primary Health Care, Obesity, Overweight, Digital Health, Artificial Intelligence, User-Computer Interface, Mobile Applications, Electronic Health Records

## Abstract

**Objective:** To evaluate the perceived usability, acceptability, and user experience (rather than the clinical effectiveness) of Boora, an AI-assisted, human-supervised digital platform prototype for longitudinal overweight and obesity care, among users and health professionals in Brazilian primary care.

**Design:** Convergent mixed-methods formative evaluation. Perceived usability was measured with the System Usability Scale (SUS) and summarised descriptively; semi-structured interviews conducted after hands-on use were analysed with codebook thematic analysis (Braun and Clarke); the two strands were integrated through a joint display. Qualitative reporting followed the Consolidated Criteria for Reporting Qualitative Research (COREQ).

**Setting:** Primary health care network of Ananindeua, Pará, within the Brazilian Unified Health System (January to February 2026).

**Participants:** Fifteen adults with overweight or obesity (BMI at least 25 kg/m2, confirmed via electronic health records) who used the patient application on their own smartphones for 24 hours, and eight primary care professionals (nurses, physicians, and a dietitian) who used the professional dashboard for approximately 20 minutes on predefined tasks with synthetic data.

**Main outcome measures:** SUS scores and qualitative themes addressing usability, acceptability, perceived usefulness, barriers, and perceived clinical and workflow fit.

**Results:** Boora showed good perceived usability in both cohorts (users mean 76.5, SD 10.3; professionals mean 77.5, SD 4.6; both above the SUS normative average of 68). Four themes emerged per cohort. Users valued an accessible interface and visible progress but described daily logging burden, fragile anticipated engagement, and digital-literacy and accessibility barriers. Professionals valued a clear interface and the prospect of panel-managed, proactive follow-up, while requiring training, AI governance, protected time, and interoperability with the national record. Integration indicated that the disengagement users anticipated was the risk professionals perceived the dashboard could help identify, whereas the educational AI assistant was the weakest and most ambiguous component for both groups.

**Conclusions:** Boora was perceived as usable and acceptable, with perceived value concentrated in human-supervised, longitudinal follow-up rather than autonomous self-tracking or AI advice. These findings concern perceived usability and acceptability, not clinical effectiveness or sustained engagement. Real-world adoption would depend on accessibility refinements, electronic-record integration, and clear AI governance aligned with the principles of Brazil’s proposed risk-based AI framework and the LGPD.

**Key points:** *What is already known on this topic:*

- Mobile and web applications can support weight management, but user engagement tends to decline over time without structured professional support, and most evidence comes from high-income countries.
- Autonomous AI advice in primary care can be unsafe or discordant with clinical guidelines, underscoring the need for human oversight and auditable systems.

*What this study adds:*

- An AI-assisted, human-supervised platform prototype pairing a patient application with a professional dashboard was perceived as usable and acceptable by both patients and primary care professionals in the Brazilian public health system, based on brief hands-on use.
- Mixed-methods integration indicated that the disengagement users anticipated corresponded to the risk professionals perceived the dashboard could help identify, locating the platform’s perceived value in human-supervised longitudinal follow-up; the educational AI assistant was its weakest component.

*How this study might affect research, practice or policy:*

- It supports designing AI in primary care obesity tools as auditable, supervised support rather than autonomous advice, consistent with the principles of Brazil’s proposed risk-based AI framework.
- Participants identified accessibility for low-literacy users, interoperability with the national electronic health record, protected time, and training as potential prerequisites for adoption, which warrant longitudinal, real-world evaluation.

## 1. Introduction

Overweight and obesity are major global public health challenges that increase health-system burden and impair quality of life^1^. By 2030, more than half of the global adult population is projected to have an elevated body mass index^1,2^. In Brazil, approximately 68% of adults already live with excess weight, which is strongly associated with noncommunicable diseases^2^.

Within the Brazilian Unified Health System, primary health care (PHC) coordinates the management of chronic conditions. However, longitudinal care for overweight and obesity remains challenging because of high service demand, limited consultation time, and fragmented care processes with inconsistent implementation of clinical protocols^3–5^.

Digital health interventions offer scalable strategies for weight management, and mobile and web applications can support weight reduction and behavioural change^6^. Nevertheless, engagement commonly declines without structured professional support^6,7^, and many applications remain poorly integrated with clinical workflows and electronic health records^8^. Evidence is also geographically concentrated: more than 93% of primary studies on digital weight-management interventions have been conducted in high-income countries^6^.

The incorporation of artificial intelligence (AI) introduces additional safety and ethical concerns. Automated systems may generate unsafe or clinically inappropriate recommendations^9–12^, and a Brazilian in silico evaluation found that large language models achieved a maximum adherence of 61.1% to national primary care guidelines for overweight and obesity^13^. These findings support professional oversight, continuous evaluation, and escalation to human care rather than autonomous clinical guidance.

Integrated platforms linking user-generated data, professional oversight, and secure communication remain relatively uncommon in PHC. To address this gap, Boora was developed as a digital platform for monitoring adults with overweight or obesity. It combines a patient-facing progressive web application for self-monitoring, education, and communication with a professional dashboard that organises user-generated data for follow-up. Designed around privacy-by-design principles, the platform uses AI as a bounded, auditable support tool while preserving human clinical decision-making.

The study was informed by a socio-technical perspective, recognising that the use of digital health technologies depends on interactions among technological infrastructure, users, and organisational workflows. Accordingly, Boora was evaluated not only in terms of interface usability but also regarding its perceived fit with PHC routines and constraints.

This study aimed to evaluate the perceived usability, acceptability, and user experience of the Boora prototype among users and primary care professionals.

## 2. Materials and Methods

### 2.1. Study design

This convergent mixed-methods study evaluated the usability, acceptability, and user experience of the Boora prototype. Quantitative and qualitative data were collected concurrently, analysed independently, and integrated during interpretation. In this article, “user” refers to interaction with the platform, whereas “patient” refers to the person receiving care. Qualitative reporting followed the Consolidated Criteria for Reporting Qualitative Research (COREQ)^14^, and methodological quality was appraised as a reporting aid using the Mixed Methods Appraisal Tool (MMAT), version 2018^15^, without computing an overall quality score. Completed instruments are provided in Appendices A and B.

In this study, acceptability refers to participants’ overall approval of, and willingness to use, the prototype as expressed in the interviews, distinguished from usability (ease of use, captured by the SUS), perceived usefulness (expected benefit), and intention to use (anticipated future use); these constructs were treated as related but separate in the coding framework.

### 2.2. The Boora platform

#### 2.2.1. System architecture and development methodology

FFSC developed the prototype across successive versions using manual programming with selective large language model assistance for interface scaffolding, style refinement, and troubleshooting, while retaining responsibility for the architecture and core logic. Development drew on clinical experience in PHC weight management and established design and usability principles^16^. No formal patient or public involvement occurred during development; this study was the first structured evaluation with prospective users and professionals. The front end used JavaScript, React, Vite, and Tailwind CSS, and the back end used Firebase, Cloud Firestore, and Cloud Functions. Package versions and the deployment date of the evaluated build are reported in Supplementary File S1.

#### 2.2.2. User and professional interfaces

Boora comprises two integrated environments on a unified database (Figure 1). The patient-facing Progressive Web App (PWA) avoids app-store installation and supports cross-device access. It enables weight and routine logging, goal tracking, access to meal plan documents, and tailored education through a simple, high-contrast interface (Figure 2). The Professional Web Dashboard organises user-generated data into longitudinal views, including weight trajectories, adherence indicators, and alerts to support follow-up (Figure 3). The dashboard’s adherence indicators and risk flags (for example, dropout risk and goal risk) were generated by deterministic, rule-based thresholds applied to logged data (for example, time since the last weight entry), not by trained or validated predictive models; no machine-learning prediction of disengagement was developed or tested.

**Figure 1.**
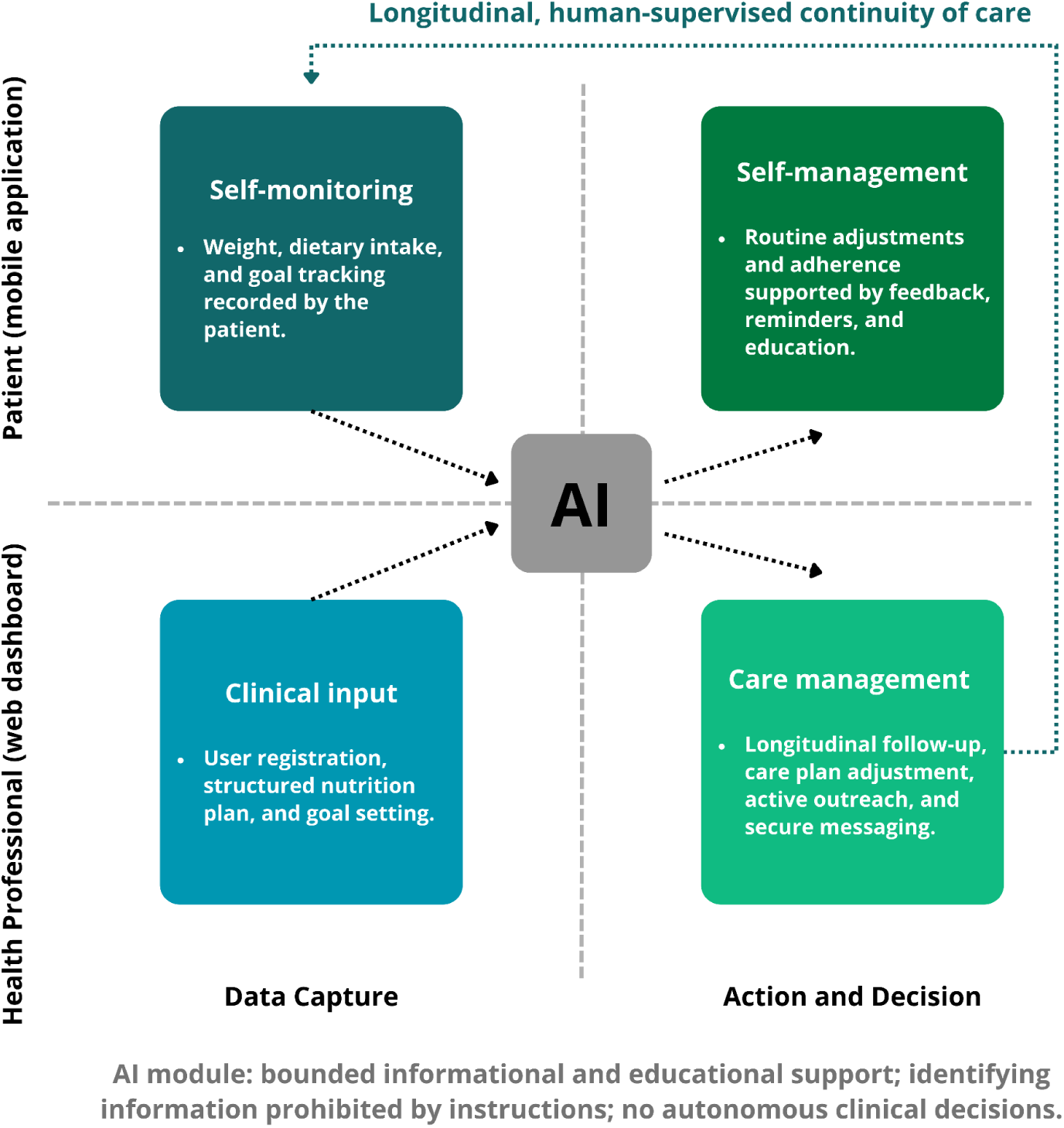
The Boora ecosystem for longitudinal overweight and obesity care in PHC. The platform pairs a patient-facing progressive web application with a professional web dashboard across two functional domains: data capture and action and decision support. Users record self-monitoring data and access feedback, reminders, educational content, and secure communication, while professionals provide clinical input and may use longitudinal data to support follow-up, care-plan adjustment, active outreach, and secure messaging. An artificial intelligence module supports selected informational and educational functions under human oversight. It does not make autonomous clinical decisions, and clinical input, data storage, monitoring, and care-management actions remain the responsibility of the application, database, and healthcare professionals. In the patient-facing assistant, users were instructed not to enter identifying information, but no automated detection, refusal, masking, or redaction of identifiers was implemented. The outer loop represents the intended human-supervised continuity of care linking user self-monitoring to professional follow-up. In this study, the patient application and professional dashboard were functional prototypes, the dashboard assistant shown to professionals was a simulated interface, and interoperability with the national electronic health record was conceptual and not implemented.

**Figure 2.**
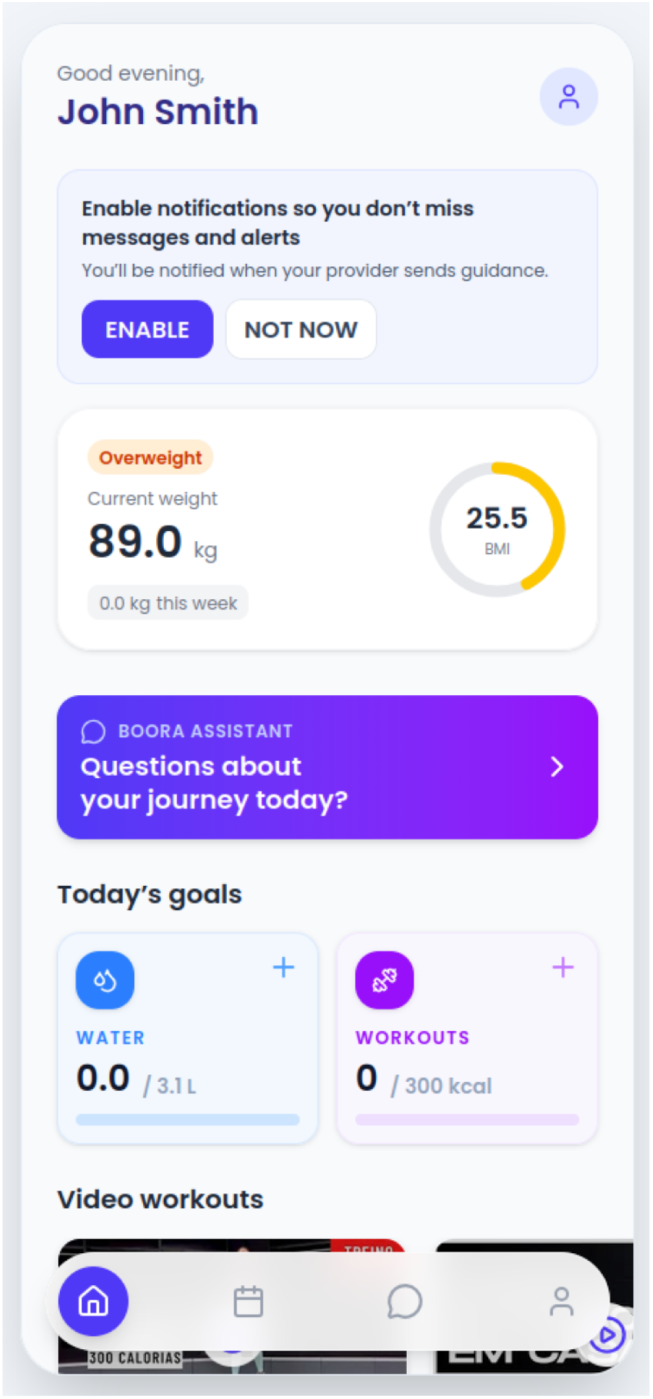
Representative screen of the Boora patient application. The home view displays current weight and body mass index, access to the educational conversational assistant, and daily self-management goals. All data are synthetic, and interface labels were rendered in English for publication.

**Figure 3.**
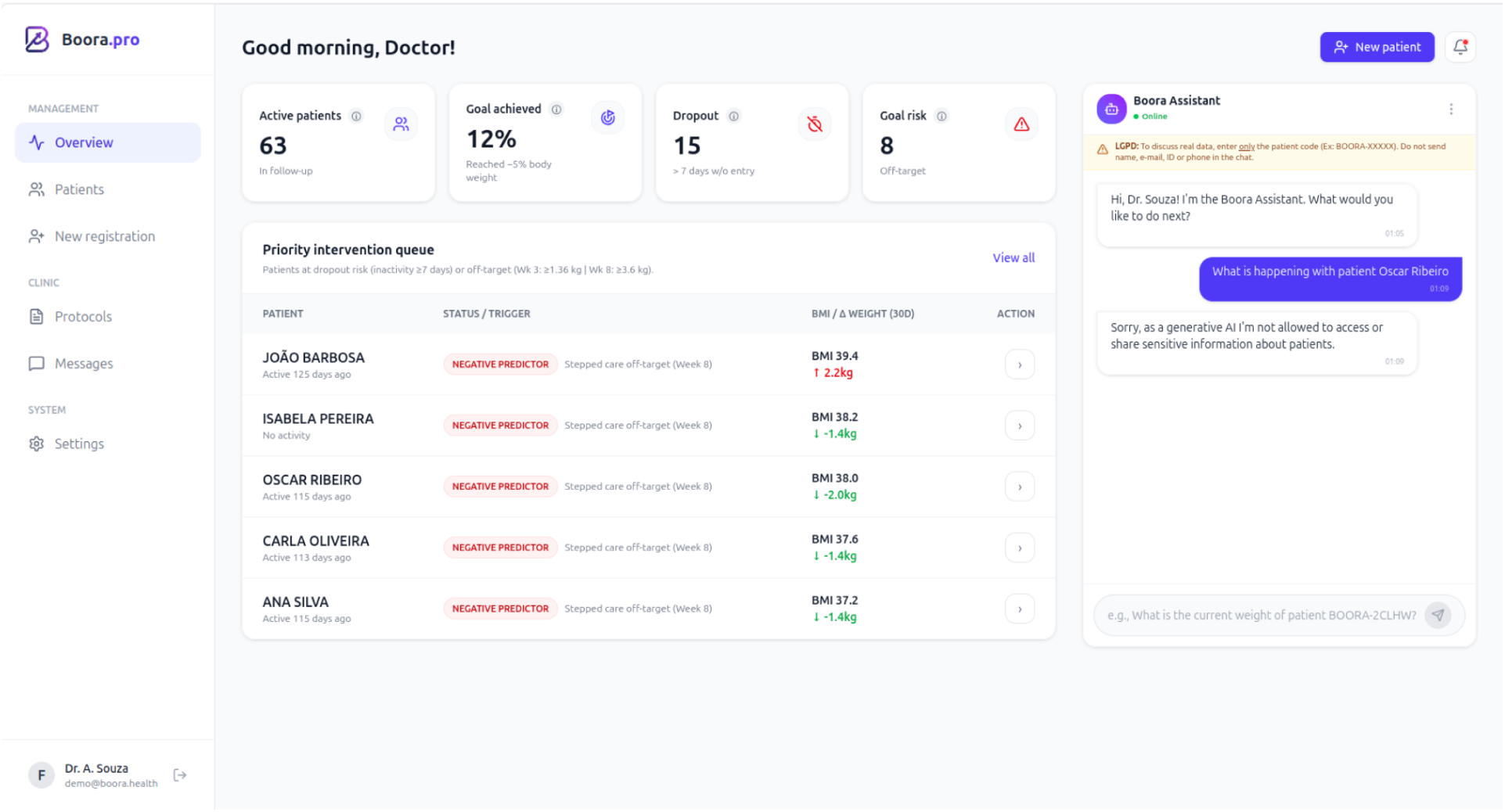
Representative screen of the Boora professional dashboard. The dashboard displays the active-patient panel, deterministic dropout- and goal-risk flags, the resulting priority intervention queue, and a simulated interaction with the embedded educational assistant. In the example, a patient is referenced by a pseudonymous code, and the assistant follows its instruction not to disclose identifying information. This illustrative interaction does not, by itself, demonstrate systematic privacy protection. All data and patient names are synthetic, and interface labels were rendered in English for publication.

#### 2.2.3. Data security, privacy, and ethical AI integration

Data protection was incorporated as a core design principle, in line with Privacy by Design and the Brazilian General Data Protection Law (LGPD). Firebase Authentication and role-based access control restricted data visibility by permission, sensitive application logic was processed through server-side Cloud Functions, and records were linked to pseudonymous identifiers rather than participant names. Required data were limited to authentication credentials and the sociodemographic and anthropometric information needed for platform operation; dietary logs, activity records, goals, free-text notes, and assistant interactions were optional.

Research data were stored in a Firebase project configured in the “southamerica-east1” region (São Paulo, Brazil) and encrypted in transit and at rest through provider-managed encryption. Vercel hosted the web interface but was not the primary data repository. Direct database access was restricted to authorised research and technical personnel, although the prototype did not include a comprehensive clinical audit trail. Data categories, access controls, retention, and assistant data flows are detailed in Supplementary File S1.

AI features were limited to informational triage and educational support and did not make autonomous clinical decisions. As no independent privacy or security assessment was conducted, the platform is described as designed to incorporate privacy-by-design and LGPD principles, rather than as demonstrating certified compliance.

#### 2.2.4. Conversational AI features

The platform included distinct patient- and professional-facing conversational features. The patient assistant accessed Google Gemini 2.0 Flash-Lite through the Google Generative Language API; no local model, fine-tuning, or project-specific training was used. A fixed Brazilian Portuguese guardrail instruction was prepended to the 12 most recent messages (Supplementary File S2), restricting responses to educational support on healthy habits and weight management, prohibiting diagnosis and medication advice, and directing users reporting warning signs to in-person or urgent care (Figure 4). The endpoint had no database access, retrieval-augmented generation, web search, or functions for writing records or initiating clinical actions. In the evaluated version, the professional dashboard used a simulated interface; a separate authenticated backend, described in Supplementary File S2, used pseudonymous patient codes, restricted Firestore snapshots, and automated redaction of selected identifiers. The study assessed perceptions, not model accuracy or clinical safety.

**Figure 4.**
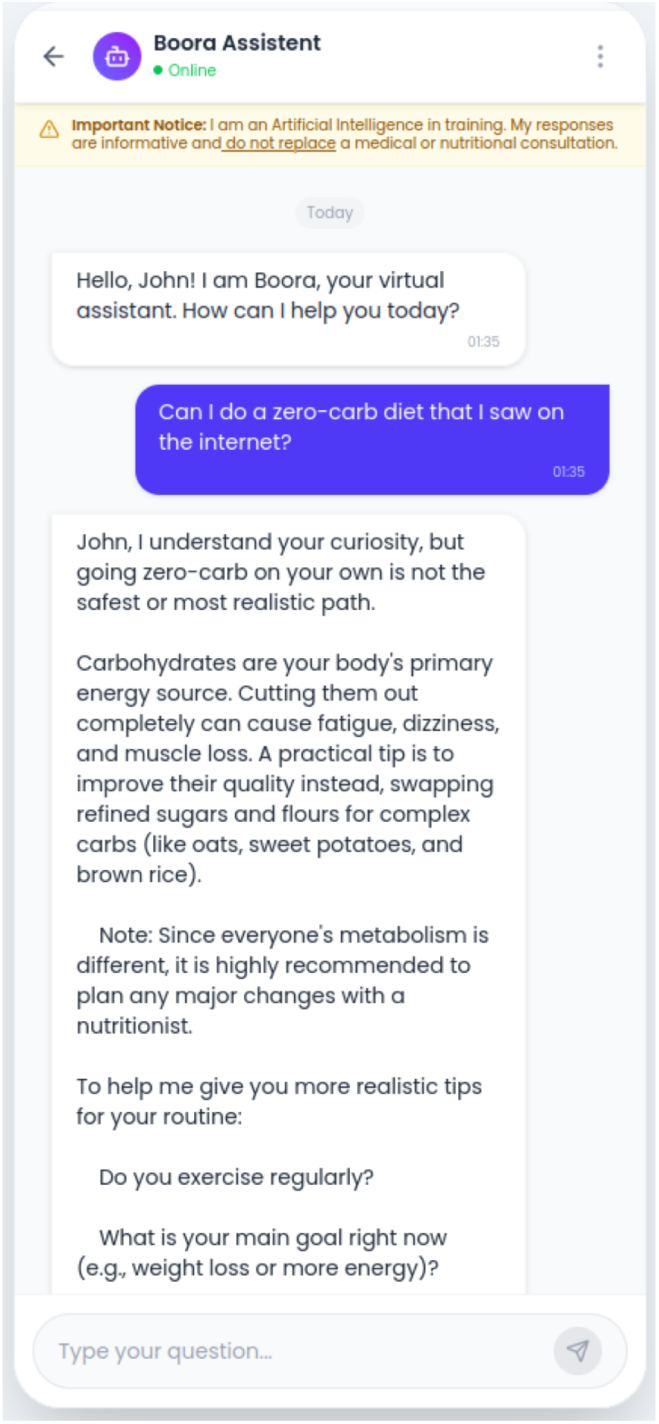
Representative patient-facing interaction with the Boora educational assistant. In this illustrative example, the user asks about adopting a zero-carbohydrate diet, and the assistant discourages the restrictive practice, provides brief general guidance, and recommends professional follow-up. The example demonstrates the assistant’s intended bounded educational behaviour but does not, by itself, establish systematic clinical accuracy or safety. Interface labels were rendered in English for publication.

### 2.3. Participants and setting

The study was conducted in the PHC network of Ananindeua, Pará, Brazil, between January and February 2026, with two participant cohorts. The user cohort comprised 15 adults aged ≥18 years with overweight or obesity (BMI ≥25 kg/m^2^) registered at a local Family Health Unit. Eligibility was verified through electronic health records; self-reported weight and height were used only for descriptive characterisation. Participants were required to own an internet-enabled smartphone and operate its basic functions. Digital literacy was not formally assessed, and assistance from a relative was permitted. Pregnancy, severe psychiatric disorders, and cognitive impairment precluding informed consent or platform use were exclusion criteria. The smartphone requirement excluded adults with the greatest digital vulnerability.

The professional cohort comprised eight physicians, nurses, and a dietitian working in municipal PHC, selected to represent different clinical perspectives and multidisciplinary workflows.

Sample sizes were pragmatically defined for a formative evaluation. Small samples are conventional in early prototype usability studies^17–19^, while qualitative sufficiency was guided by information power^20^, considering the focused research aim, specific information-rich cohorts, and structured, theory-informed analysis. Saturation was not used as a stopping criterion.

Potential users were identified from an authorised extract of the *Prontuário Eletrônico do Cidadão*, the electronic health record component of Brazil’s e-SUS Primary Health Care strategy (PEC/e-SUS APS). The extract was filtered for BMI ≥25 kg/m^2^, with pregnant individuals excluded, and records were ordered by the date of the most recent consultation. Records were reviewed consecutively, and the first 15 eligible users were invited by FFSC; all accepted, with no refusals or subsequent exclusions. Professionals were identified through an alphabetical list of municipal primary care units obtained from the National Registry of Health Establishments and were contacted sequentially by email. Of the 11 professionals contacted, three did not respond within seven working days and eight enrolled. The recruitment flow is presented in Supplementary File S3.

### 2.4. Data collection

Data collection combined brief hands-on prototype exposure, the SUS, and semi-structured interviews. Fifteen user interviews were conducted in person in a private room at the participating primary care unit, and eight professional interviews by secure videoconference. Users completed a sociodemographic questionnaire, activated the PWA on their smartphones, and used it for 24 hours; assistance from a relative was permitted and recorded. In a second session, they completed the Brazilian Portuguese SUS^21^ and an individual interview. Professionals completed sociodemographic and professional questionnaires, used the dashboard for approximately 20 minutes to perform predefined tasks with synthetic data (Supplementary File S4), completed the SUS, and were interviewed. No professional accessed data from the user cohort. Objective task-performance metrics and platform-use logs were not collected.

Interview guides addressed usability, perceived usefulness, barriers, facilitators, anticipated clinical utility, and integration into care routines (Supplementary Files S5 and S6) and were not formally pilot-tested. User interviews lasted a mean of 19.8 minutes (range 12-31) and professional interviews 28.4 minutes (range 19-43), excluding prototype exposure and SUS completion. No repeat interviews were conducted. Coded data and the codebook were managed in spreadsheets.

All interviews were audio-recorded and conducted by FFSC, a male family physician and master’s student who developed the platform and had previously worked as the users’ family physician, although not at the time of the interviews. FFSC had postgraduate training in qualitative methods, interviewing, thematic analysis, and COREQ reporting. Participants knew that he was a master’s candidate and that the study formed part of his research. Interviews were conducted individually with no one else present. FFSC transcribed the recordings verbatim and checked each transcript against the audio. No field notes, transcript return, or member checking were undertaken; analytical notes were recorded during familiarisation. Standardised scripts and explicit encouragement of critical feedback were used to mitigate reflexivity and social desirability bias.

### 2.5. Data analysis

Quantitative and qualitative data were analysed independently and then integrated. SUS scores were summarised by cohort using mean, standard deviation, median, interquartile range, and minimum-maximum values. Standard scoring converted the ten items to a 0-100 scale; all items were complete. Scores were interpreted descriptively as below average (<68), good (68-80.3), or excellent (>80.3), with 68 treated as a normative average rather than an individual pass-fail threshold^22–25^. No inferential between-group comparison was performed because of the small samples and different interfaces and exposure conditions.

Complete Portuguese-language transcripts were analysed using codebook thematic analysis^26,27^, following Braun and Clarke’s six phases within a structured approach combining deductive sensitising concepts with inductively generated codes. Analysis addressed usability, perceived usefulness, barriers, facilitators, anticipated clinical utility, and integration with workflows, workload, roles, and health-system structures. Reporting followed COREQ^14^.

FFSC coded all 15 user and eight professional transcripts. Before codebook consolidation, CPBA independently coded a maximum-variation subsample of three user and three professional interviews representing contrasting experiences of usability, accessibility, workload, perceived clinical value, and AI governance. CPBA holds a PhD, supervises qualitative and mixed-methods research, and contributed to the interview guides and analytical development.

The authors compared codes by semantic meaning, reconciled differences through discussion, and consolidated a shared codebook. FFSC then applied it across the full dataset, refining definitions iteratively. Codes were grouped into categories, and themes were reviewed against coded extracts and complete transcripts for coherence, distinctiveness, and convergence or divergence between cohorts. Trustworthiness was supported by collaborative codebook development and an audit trail; inter-rater reliability was not calculated because secondary coding aimed at interpretive refinement rather than agreement measurement. The codebook and code-category-theme hierarchy are provided in Supplementary File S7. Quotations were translated after theme development and reviewed by both authors for semantic and contextual equivalence; formal back-translation was not performed.

SUS findings and qualitative themes were integrated through a joint display^28^ to identify convergence, complementarity, and divergence and derive meta-inferences about perceived usability, acceptability, anticipated clinical utility, and implementation conditions.

### 2.6. Ethical considerations

The study complied with the Declaration of Helsinki and Brazilian National Health Council Resolutions No. 466/2012 and No. 510/2016. The protocol was approved by the Research Ethics Committee of the Institute of Health Sciences, Federal University of Pará (CAAE No. 91547525.4.0000.0018; Approval Opinion No. 8.088.382), and all participants provided written or electronic informed consent.

Eligibility was verified through electronic health records by authorised personnel. Study data were pseudonymised using alphanumeric codes, with code-to-identity keys accessible only to authorised personnel, and will be securely destroyed after the five-year retention period established in the protocol.

Participants were informed that the 24-hour exposure was solely for usability and user-experience assessment and did not replace usual care, professional advice, or emergency services. Platform entries were not monitored clinically in real time, and no individual feedback was provided through the platform; data were reviewed only after the exposure period to characterise use and inform the interviews. No platform-related clinical or safety events were reported. Relatives could assist with installation and data entry, with the possibility of viewing the information entered.

### 2.7. Patient and public involvement

Patients and members of the public were not formally involved in defining the research question or study design. However, the platform was developed to address practical needs observed in primary care obesity follow-up, and users participated in usability testing and provided feedback on interface clarity, perceived usefulness, and participation burden. Findings will be disseminated through academic outputs and may inform future participatory refinement of the platform.

## 3. Results

### 3.1. Participant characteristics

All 15 users and 8 professionals who enrolled completed all procedures, with no withdrawals. Users had a mean age of 40.6 years (SD 12.4; range 24 to 60), and 9 (60.0%) were women (Table 1, Panel A). All had overweight or obesity confirmed through electronic records; self-reported measurements produced a mean BMI of 32.1 kg/m^2^ (SD 3.5). Six (40.0%) reported incomplete or completed Brazilian fundamental education, corresponding to primary and lower secondary education, and 14 (93.3%) had a monthly family income of up to three minimum wages. Although all owned an internet-enabled smartphone, use frequency varied, and 12 (80.0%) had never used a health application or had discontinued previous use.

**Table 1.** Characteristics of participants.

**Panel A. Users (n = 15)**
| Characteristic | Value |
| --- | --- |
| Age, years, mean (SD); range | 40.6 (12.4); 24 to 60 |
| Women, n (%) | 9 (60.0) |
| BMI, kg/m<sup>2</sup>, mean (SD); range | 32.1 (3.5); 27.9 to 37.4 |
| Overweight (25 to 29.9), n (%) | 5 (33.3) |
| Obesity class I (30 to 34.9), n (%) | 5 (33.3) |
| Obesity class II (35 to 39.9), n (%) | 5 (33.3) |
| Obesity class III ($\geq 40$), n (%) | 0 (0.0) |
| Fundamental education or less (complete or incomplete), n (%) | 6 (40.0) |
| Family income up to 1 minimum wage, n (%) | 5 (33.3) |
| Family income 1 to 3 minimum wages, n (%) | 9 (60.0) |
| Family income 3 to 5 minimum wages, n (%) | 1 (6.7) |
| Owns internet-enabled smartphone, n (%) | 15 (100) |
| Smartphone use every day, many times, n (%) | 6 (40.0) |
| Smartphone use every day, a few times, n (%) | 5 (33.3) |
| Smartphone use a few times per week, n (%) | 4 (26.7) |
| Prior health-app use, never / discontinued / current, n | 7 / 5 / 3 |

**Panel B. Health professionals (n = 8)**
| Characteristic | Value |
| --- | --- |
| Age, years, mean (SD); range | 39.9 (6.3); 29 to 48 |
| Women, n (%) | 5 (62.5) |
| Profession, nurse / physician / dietitian, n | 4 / 3 / 1 |
| Years since graduation, mean (SD); range | 14.8 (6.2); 5 to 23 |
| Primary care experience over 6 years, n (%) | 7 (87.5) |
| Holds a postgraduate qualification, n (%) | 8 (100) |
| Digital-technology familiarity, high / moderate, n | 4 / 4 |
| Recommends health apps to patients, frequently / rarely, n | 4 / 4 |

*BMI, body mass index; SD, standard deviation. Eligibility (BMI at least 25 kg/m<sup>2</sup>) was confirmed via electronic health records; the weight and height used for the BMI values and categories shown here were self-reported and are presented for descriptive characterisation only. Self-reported anthropometry may misclassify obesity class. BMI categories follow World Health Organization cut-offs.*

The professional cohort (Table 1, Panel B) comprised 4 nurses, 3 physicians, and 1 dietitian, with a mean age of 39.9 years (SD 6.3) and a mean of 14.8 years since graduation (SD 6.2; range 5 to 23). All 8 professionals held a postgraduate qualification, most commonly in Family Health (4) or Public Health (3), with one in Clinical Nutrition, and 7 (87.5%) had worked in primary care for more than six years. Self-rated familiarity with digital technologies was high in 4 (50.0%) and moderate in 4 (50.0%), and the practice of recommending health applications to patients was evenly split between frequent (4) and rare (4).

### 3.2. Perceived usability assessed using the SUS

All 15 users and all 8 health professionals completed the SUS. Overall, Boora showed good perceived usability in both cohorts, with mean scores above the normative average of 68. Among users, the mean SUS score was 76.5 (SD 10.3; median 75.0, IQR 68.8 to 78.8; range 67.5 to 100). Among health professionals, the mean SUS score was 77.5 (SD 4.6; median 78.8, IQR 74.4 to 80.6; range 70 to 82.5). Scores were more homogeneous among professionals, whereas users displayed greater dispersion (Table 2).

**Table 2.** SUS scores among users and health professionals.

| Statistic | Users (n = 15) | Health professionals (n = 8) |
| --- | --- | --- |
| Mean (SD) | 76.5 (10.3) | 77.5 (4.6) |
| Median (IQR) | 75.0 (68.8 to 78.8) | 78.8 (74.4 to 80.6) |
| Minimum to maximum | 67.5 to 100 | 70 to 82.5 |
| Excellent, n (%) | 4 (26.7) | 2 (25.0) |
| Good, n (%) | 7 (46.7) | 6 (75.0) |
| OK, n (%) | 4 (26.7) | 0 (0) |
| Score at or above 68, n (%) | 11 (73.3) | 8 (100) |

*SD, standard deviation; IQR, interquartile range. Descriptive bands: below average (< 68), good (68 to 80.3), and excellent (> 80.3). A SUS score of 68 represents the normative average and is not an individual pass or fail threshold.*

The mean scores for both groups fell within the good band. Four users (26.7%) and two professionals (25.0%) reached the excellent band, while the four lowest user scores (67.5) fell just below the normative average of 68 (Table 2). Internal consistency was high among users (Cronbach alpha 0.94) and acceptable among professionals (0.71). At item level, the lowest-scoring items in both cohorts concerned the need for technical support and the amount of learning required before use, and among users also confidence in use (Supplementary File S8). User and professional scores are reported side by side for description only and are not directly comparable, because the cohorts used different interfaces, performed different tasks, and had markedly different exposure durations.

### 3.3. Qualitative findings

Codebook thematic analysis of 15 user interviews (180 coded excerpts) and 8 professional interviews (128 coded excerpts) generated four themes for each cohort. User themes concerned onboarding and logging burden, the AI assistant, engagement, and digital inclusion (U-T1 to U-T4); professional themes concerned interface fit, panel-managed care, safe implementation, and systemic integration (P-T1 to P-T4). These labels are used for cross-reference in Table 3 and Supplementary File S7.

**Table 3.**
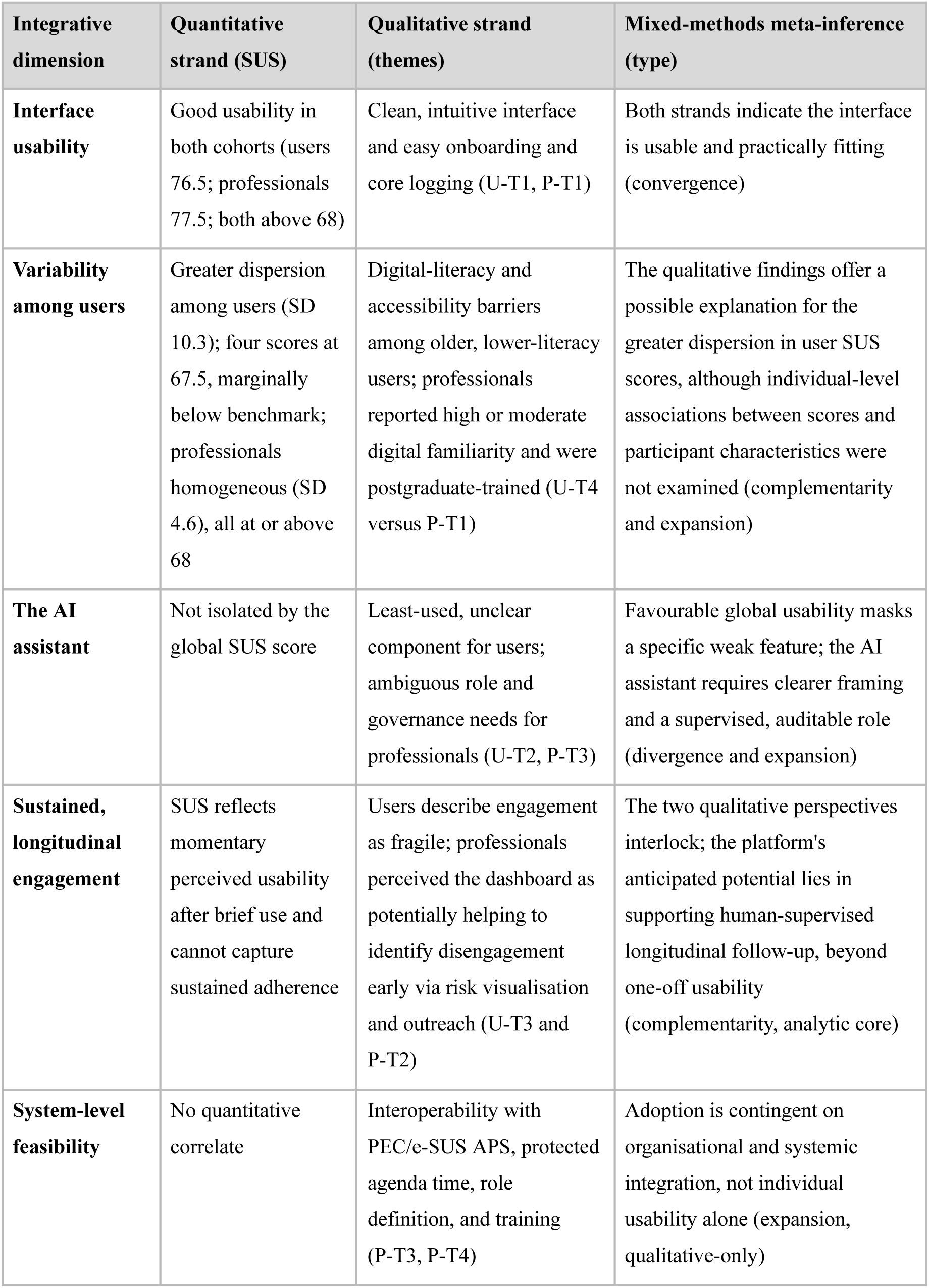
Joint display of SUS findings and qualitative themes.

Quotations were translated from Brazilian Portuguese by the authors; ellipses indicate omitted speech and square brackets denote non-verbal expressions. PEC refers to the Prontuário Eletrônico do Cidadão, the electronic health record component of Brazil’s nationally deployed PEC/e-SUS APS strategy.

#### 3.3.1. Users’ experience

Accessible onboarding alongside effortful daily logging. Users described a positive first impression and a low barrier to entry. The interface was characterised as clean, modern, and easy to navigate, and core actions such as recording weight or water intake were experienced as immediate and intuitive. As one younger participant summarised, “I found the experience really interesting. The app has a modern feel… It looked clean and was very easy to navigate right from the start” (BOORA-CP011). However, dietary logging emerged as the principal source of friction: participants found it laborious, reported difficulty locating regional foods, and described the screen as containing too many fields. A participant with no prior app experience recalled, “When it came to the food part, I understood nothing [laughs]… I thought there was too much to click… there are too many functions there for my head” (BOORA-CP009). This contrast between intuitive core tracking and burdensome meal logging underpinned concerns about sustained use.

The educational AI assistant emerged as the least adopted and most ambiguously understood feature. Users commonly reported not engaging with it because its purpose was unclear or because they were still learning other functions: “I saw there was a chat there, but… I thought it was better not to even touch it, so as not to confuse myself more. I didn’t really understand what it was for” (BOORA-CP002). A minority valued it as a source of quick guidance: “I found the AI chat really interesting. It clears up some doubts, gives you a bit of light” (BOORA-CP012). One digitally fluent user preferred a general-purpose tool she already used and returned to the platform’s logging functions (BOORA-CP014). The assistant was therefore perceived as a non-essential component whose usefulness depended on clearer explanation of its role.

Self-monitoring was an important source of motivation, although participants anticipated that engagement could weaken over time. Visual tracking of weight and progress made their efforts more tangible, whereas repetitive data entry, forgetfulness, and difficulties with digital technologies created a risk of discontinuation. One participant captured this tension: “What motivates me is seeing that I’m meeting the goals, getting close to the target. Now, what discourages me is having to keep weighing food or measuring everything I eat… if it’s too much, you end up stopping, just like happened with other [apps] I’ve had before” (BOORA-CP012). Professional oversight was also anticipated as a motivator: “What really motivates me is knowing there’s someone on the other side following along… knowing the doctor will see what I’m doing and will give me feedback afterwards” (BOORA-CP007). This statement reflects the anticipated value of professional oversight rather than monitoring during the study, as platform data were not reviewed in real time. Overall, engagement appeared to depend more on visible progress and perceived professional connection than on self-tracking alone.

Digital literacy and accessibility emerged as conditions for inclusive and autonomous use. Although most participants navigated the platform, some with limited digital experience depended on relatives for installation or data entry: “I can’t use it without her by my side. If I’m on my own, I don’t know where to click, I can’t see the picture properly… I needed her help the whole time” (BOORA-CP010). Participants also requested larger fonts and touch targets and simpler language: “I found the letters a bit too tiny… the boxes and the letters could be bigger to make it easier on the eyes” (BOORA-CP002). Thus, broader inclusion would require accessibility refinements and support during initial use.

#### 3.3.2. Health professionals’ experience

Professionals anticipated that the platform would fit clinical practice because of its clear, objective, and visually uncluttered interface. These characteristics were linked to the time constraints of primary care, where information must be interpreted rapidly. The hierarchical presentation was perceived as facilitating clinical reasoning: “Objectivity is the strong point. In the clinical setting, we have little time to interpret complex data… the panel presenting information in a hierarchical, clean way lets clinical reasoning be faster. We don’t get lost in unnecessary menus” (BOORA-P008). Risk foregrounding was viewed as supporting prioritisation without adding visual complexity: “Boora highlighting what is a priority, like the risk alert, without cluttering the screen, makes it much more intuitive to use” (BOORA-P002). Perceived suitability was commonly conditioned on an initial period of familiarisation.

Perceived clinical utility was most strongly associated with the platform’s potential to support longitudinal, panel-managed care. Professionals envisaged a shift from episodic follow-up toward proactive management of a care panel, with risk and adherence indicators potentially supporting earlier outreach. Several interpreted this as work organisation rather than additional workload: “Today in primary care, follow-up is very passive; we wait for the patient to miss an appointment before we notice. With this panel we can be more proactive… this isn’t an overload, it ends up being work intelligence, because it prevents the patient from getting worse” (BOORA-P006). The panel was also perceived as supporting coordination across a caseload: “In primary care we often lose sight of the patient in the territory. Having this panel helps to manage not just the individual but the group… we stop firefighting and start doing case management in a more planned way” (BOORA-P008). These accounts represent anticipated utility, as the dashboard was evaluated through simulated tasks rather than routine clinical use.

Professionals identified training, AI and messaging governance, role definition, and workload management as conditions for safe implementation. The AI assistant’s function was a recurrent source of ambiguity: “…the exact function of the AI assistant, is it an assistant for case discussion or for general questions?” (BOORA-P002). Direct messaging was likewise perceived as requiring protocols to protect data and prevent indiscriminate use. Workload concerns centred on organisation and protected time: “…it needs organisation so as not to create overload… if we define the roles well, who monitors dropout and who delivers the clinical intervention, it optimises a lot. The risk is that it just becomes one more thing to open, without protected time in the schedule” (BOORA-P004). Participants suggested brief in-system tutorials and prior training before routine adoption.

Systemic integration was viewed as a basis for continuity of care, with interoperability with PEC/e-SUS APS emerging as a key perceived condition for sustained adoption. Integration was expected to reduce duplicate data entry and provide a more complete view of the patient: “If the weight data and the food diary the patient records in the app went straight into the medical record, it would avoid the rework of opening two screens… I would have the real history of what the patient is doing outside the office available the moment I open their file in the e-SUS. That greatly improves the safety of what I’m prescribing” (BOORA-P001). It was also linked to multiprofessional continuity and the legitimacy of an official record: “If I record a reminder or a goal agreement in Boora and it appears in the record, any other colleague who sees this patient later will know exactly where we left off… that strengthens the bond and the patient’s trust in the service” (BOORA-P006). Without interoperability, professionals anticipated that the platform could remain isolated from existing workflows.

### 3.4. Integration of quantitative and qualitative findings

Following a convergent mixed-methods logic, SUS results and qualitative themes were integrated through a joint display (Table 3) to identify convergence, complementarity, expansion, and divergence and to derive meta-inferences. User themes are identified as U-T1 to U-T4 and professional themes as P-T1 to P-T4, as defined in Section 3.3 and Supplementary File S7.

Overall, integration indicated favourable perceived interface usability while identifying feature-specific, engagement-related, and organisational limitations not captured by the global SUS scores. These meta-inferences were interpretive rather than statistical, and no causal or implementation effects were assessed.

## 4. Discussion

Boora showed favourable perceived usability in both cohorts, with mean SUS scores above the normative average of 68, but the integrated findings qualified this overall result. Users described logging burden, digital-accessibility barriers, and anticipated difficulties sustaining engagement, whereas professionals perceived that panel-based monitoring could potentially support earlier identification and outreach. The educational AI assistant was the least clearly understood component, showing that favourable global usability may coexist with feature-specific weaknesses. Professionals also identified interoperability, protected time, role definition, training, and AI governance as perceived conditions for adoption. Together, these findings position Boora as a potential human-supervised follow-up tool rather than a stand-alone self-management or autonomous AI intervention.

These findings are consistent with evidence that mobile and web applications can support weight management and behavioural change, while sustained engagement commonly declines without structured professional support^6,7^. In the present study, visible progress encouraged self-management, but repetitive logging and routine constraints were perceived as potential drivers of discontinuation. Professional oversight was itself anticipated as a source of motivation. This interpretation is consistent with evidence associating continuity with a clinician with improved outcomes^29^ and suggests that longitudinal relationships may be central to the sustainability of digital interventions in primary care.

The educational AI assistant was little used and ambiguously understood by both cohorts. The study assessed these perceptions rather than the assistant’s technical performance or safety. Although large language models can perform well in circumscribed tasks^30,31^, they may also generate unsafe or guideline-discordant recommendations^11,12^; an in silico evaluation by the authors found a maximum adherence of 61.1% to Brazilian primary care guidelines for overweight and obesity^13^. The present findings therefore support a bounded, human-supervised role for AI, with clear scope, escalation pathways, and governance, rather than autonomous clinical guidance.

Boora was designed to incorporate privacy-by-design and LGPD principles, although no independent privacy or security assessment was conducted. World Health Organization guidance similarly emphasises human oversight, transparency, and post-deployment auditing of health AI^32^. Brazil’s proposed risk-based AI framework remained under legislative consideration at the time of writing^33^, while enacted international frameworks, such as the European Union AI Act, classify some health-related applications according to their functions and risks^34^. Regardless of future legal classification, professionals’ requests for protocols, role definition, and data protection indicate that governance is also a condition for perceived usability and acceptability.

The findings also highlight digital equity and care coordination. Some participants with limited digital experience required family assistance and requested larger fonts, touch targets, and simpler language. These barriers are particularly relevant because digital weight-management evidence remains concentrated in high-income settings^6^, while infrastructure and digital-literacy constraints persist in developing health systems^35^. Although connectivity and electronic-record use are widespread in Brazilian primary care facilities, household access remains unequal and is often smartphone-dependent^36–38^. Professionals therefore viewed interoperability with PEC/e-SUS APS, protected time, training, and role definition as important conditions for integrating Boora into coordinated care.

This study has several limitations. It evaluated a prototype over a brief period, 24 hours for users and approximately 20 minutes of simulated tasks for professionals, using small, non-representative cohorts from a single municipality. The study therefore assessed initial perceived usability, anticipated acceptability, and perceived workflow fit rather than sustained engagement, implementation, clinical effectiveness, or effects on continuity of care. No objective performance measures or platform-use logs were collected, and the cohorts used different interfaces under different exposure conditions; their SUS scores should therefore not be directly compared. Professionals interacted with synthetic scenarios rather than using the platform in routine care, and no end-to-end patient-professional workflow, generated alert, or electronic-record integration was evaluated. The AI assistant was assessed only through participants’ perceptions; its technical performance, guardrail compliance, and clinical safety were not tested.

Additional limitations concern measurement, selection, and researcher influence. Descriptive anthropometry was self-reported, interview modes differed between cohorts, quotations were not formally back-translated, and quantitative and qualitative data were not linked at the individual level. The primary author developed the platform, had previously provided care to participating users, identified and invited them, and conducted the interviews, creating risks of reflexivity and socially desirable responses. Standardised scripts, explicit encouragement of criticism, and collaborative codebook development were used to mitigate these risks but could not eliminate them. Smartphone and internet access requirements excluded adults with the greatest digital vulnerability, and three of the 11 professionals contacted did not respond. Usability and acceptability may therefore be overestimated for the wider PHC population, and the findings should be interpreted as transferable qualitative insights rather than representative estimates.

Future studies should evaluate Boora longitudinally under routine care conditions, measuring sustained engagement, continuity of care, workflow burden, and clinically relevant outcomes. Priorities include objective usability and usage metrics, participatory accessibility refinement, evaluation among people with lower digital access, interoperability with PEC/e-SUS APS, and technical and safety assessment of the AI and messaging components under an explicit governance framework aligned with the LGPD and Brazil’s proposed risk-based AI legislation.

## 5. Conclusion

In this formative mixed-methods evaluation, Boora was perceived as usable and acceptable, and as potentially relevant to care, by both users and primary care professionals in the Brazilian Unified Health System. Its perceived value lay primarily in supporting human-supervised, longitudinal follow-up: the disengagement that users anticipated corresponded to the risk that professionals perceived the dashboard could help identify. The educational AI assistant was the least adopted component, reinforcing a design in which AI serves as auditable support rather than autonomous guidance. These findings concern perceived usability and acceptability after brief exposure, not clinical effectiveness, sustained engagement, or implementation outcomes. Whether Boora can improve continuity and outcomes will depend on accessibility refinements for low-literacy users, interoperability with the national electronic health record, protected professional time, and AI governance aligned with the LGPD and the principles of Brazil’s proposed risk-based AI framework, all of which warrant evaluation in longitudinal, real-world implementation studies.

## Declarations

### Contributors

FFSC conceived the study, designed and developed the Boora platform, collected the data, conducted the interviews, performed the primary coding and the quantitative analysis, and drafted the manuscript. CPBA contributed to the methodological design, acted as the second analyst on the strategic interview sample, critically reviewed the codebook, categories, and thematic map, supervised the analytical process, and revised the manuscript critically for important intellectual content. Both authors approved the final version and agree to be accountable for all aspects of the work. FFSC is the guarantor and accepts full responsibility for the conduct of the study, had access to the data, and controlled the decision to publish.

### Patient and public involvement

Patients and the public were not formally involved in the design or conduct of the study; the platform was developed to address practical needs in the primary care follow-up of adults with overweight or obesity, and users contributed feedback during usability testing.

### Patient consent for publication

All participants provided written or electronic informed consent to participate, data were pseudonymised (de-identified) using alphanumeric codes, and only de-identified excerpts that do not allow individual identification are reported.

### Ethics approval

This study involves human participants and was approved by the Research Ethics Committee of the Institute of Health Sciences, Federal University of Pará (UFPA), under Certificate of Presentation for Ethical Review (CAAE) No. 91547525.4.0000.0018 (Approval Opinion No. 8.088.382). Participants gave informed consent before taking part.

### Data availability statement

Data are available upon reasonable request. Due to the qualitative nature of the interview data and the risk of participant identification in a small primary care setting, full transcripts are not publicly available. De-identified excerpts supporting the findings are included in the manuscript. Aggregated SUS data and supplementary methodological materials may be made available by the corresponding author upon reasonable request, subject to ethical approval and applicable legal restrictions. The interview guides, the consolidated codebook, and item-level SUS data are provided as supplementary files.

### Supplementary Materials

The following supporting information can be downloaded at: https://doi.org/10.6084/m9.figshare.32989955, Supplementary File S1. Technical specifications, development assistance, data governance, and data flows of the Boora prototype; Supplementary File S2. Prompts and implementation details of the Boora conversational-assistant features; Supplementary File S3. Recruitment and completion flow for the user and professional cohorts; Supplementary File S4. Standardised professional-dashboard exposure tasks; Supplementary File S5. Semi-structured interview guide for professionals; Supplementary File S6. Semi-structured interview guide for user participants; Supplementary File S7. Consolidated codebook and code-category-theme hierarchy; Supplementary File S8. Item-level System Usability Scale (SUS) scores among users and health professionals.

### Use of generative artificial intelligence

OpenAI Codex was used selectively during prototype development for interface scaffolding, CSS and component refinement, code suggestions, and troubleshooting. FFSC reviewed, edited, executed, and tested all suggested code and retained responsibility for the system architecture, data model, core logic, security rules, clinical scope, and final implementation.

