## Appendix A. Consolidated Criteria for Reporting Qualitative Research (COREQ): 32-item checklist for "Boora, an AI-assisted digital platform for overweight and obesity care in Brazilian primary care: a formative mixed-methods evaluation of perceived usability and acceptability" (from another version of this preprint: version 1, 2026-07-16)

| No. | Item | Guide questions / description | Reported in |
|---|---|---|---|
| ***Domain 1: Research team and reflexivity*** | | | |
| ***Personal characteristics*** | | | |
| 1 | Interviewer / facilitator | Which author/s conducted the interview or focus group? | §2.4 (all interviews conducted by FFSC) |
| 2 | Credentials | What were the researcher's credentials? E.g. PhD, MD | §2.4 (FFSC, family physician and master's student); §2.5 (CPBA, PhD) |
| 3 | Occupation | What was their occupation at the time of the study? | §2.4; §2.5 |
| 4 | Gender | Was the researcher male or female? | §2.4 (male); §2.5 (male) |
| 5 | Experience and training | What experience or training did the researcher have? | §2.4 (postgraduate training in qualitative methods, interviewing, thematic analysis, and COREQ reporting); §2.5 (CPBA supervises qualitative and mixed-methods research) |
| ***Relationship with participants*** | | | |
| 6 | Relationship established | Was a relationship established prior to study commencement? | §2.4 (FFSC had previously been the users' family physician, not at the time of the interviews) |
| 7 | Participant knowledge of the interviewer | What did the participants know | §2.4 |

| No. | Item | Guide questions / description | Reported in |
|---|---|---|---|
| | | about the researcher? | |
| 8 | Interviewer characteristics | What characteristics were reported about the interviewer/facilitator? | §2.4; §4 (Limitations); Competing interests |
| ***Domain 2: Study design*** | | | |
| 9 | Methodological orientation and theory | What methodological orientation was stated to underpin the study? | §2.1 (convergent mixed methods; socio-technical perspective); §2.5 (codebook thematic analysis) |
| ***Participant selection*** | | | |
| 10 | Sampling | How were participants selected? | §2.3 (purposive) |
| 11 | Method of approach | How were participants approached? | §2.3 (users identified from an authorised PEC/e-SUS extract and invited by FFSC; professionals contacted by email); Supplementary File S3 |
| 12 | Sample size | How many participants were in the study? | §2.3; §3.1 (15 users; 8 professionals) |
| 13 | Non-participation | How many people refused to participate or dropped out? Reasons? | Reported. §2.3: no user refusals or exclusions after record review; 3 of 11 professionals contacted did not respond within seven working days. Supplementary File S3 |
| ***Setting*** | | | |

| No. | Item | Guide questions / description | Reported in |
|---|---|---|---|
| 14 | Setting of data collection | Where was the data collected? | §2.4 (15 user interviews in person in a private room at the participating primary care unit; 8 professional interviews by secure videoconference) |
| 15 | Presence of non-participants | Was anyone else present besides the participants and researchers? | Reported (none). §2.4: interviews conducted individually with no one else present |
| 16 | Description of sample | What are the important characteristics of the sample? | §3.1; Table 1 |
| ***Data collection*** | | | |
| 17 | Interview guide | Were questions, prompts, guides provided by the authors? Was it pilot tested? | Guides provided: Supplementary Files S5 and S6. Reported (not pilot tested): §2.4 |
| 18 | Repeat interviews | Were repeat interviews carried out? If yes, how many? | Reported (none). §2.4: no repeat interviews were conducted |
| 19 | Audio / visual recording | Did the research use audio or visual recording to collect the data? | §2.4 (all interviews audio-recorded) |
| 20 | Field notes | Were field notes made during and/or after the interview or focus group? | Reported (none). §2.4: no field notes; analytical notes recorded during familiarisation |
| 21 | Duration | What was the duration of the interviews or focus group? | §2.4 (users: mean 19.8 min, range 12–31; professionals: mean 28.4 min, range 19–43; excluding prototype exposure |

| No. | Item | Guide questions / description | Reported in |
|---|---|---|---|
| | | | and SUS completion) |
| 22 | Data saturation | Was data saturation discussed? | Reported (not used). §2.3: saturation was not used as a stopping criterion; sufficiency guided by information power † |
| 23 | Transcripts returned | Were transcripts returned to participants for comment and/or correction? | Reported (not undertaken). §2.4 |
| ***Domain 3: Analysis and findings*** | | | |
| ***Data analysis*** | | | |
| 24 | Number of data coders | How many data coders coded the data? | §2.5 (two: FFSC coded all transcripts; CPBA independently coded a subsample) |
| 25 | Description of the coding tree | Did authors provide a description of the coding tree? | Provided: Supplementary File S7 (codebook and code–category–theme hierarchy). Process described in §2.5 |
| 26 | Derivation of themes | Were themes identified in advance or derived from the data? | §2.5 (deductive sensitising concepts combined with inductively generated codes) |
| 27 | Software | What software, if applicable, was used to manage the data? | Reported (none dedicated). §2.4: coded data and codebook managed in spreadsheets |
| 28 | Participant checking | Did participants provide feedback on the findings? | Reported (not undertaken). §2.4: member checking was not undertaken |
| ***Reporting*** | | | |

| No. | Item | Guide questions / description | Reported in |
|---|---|---|---|
| 29 | Quotations presented | Were participant quotations presented to illustrate the themes / findings? Was each quotation identified? | §3.3 (quotations identified by participant code) |
| 30 | Data and findings consistent | Was there consistency between the data presented and the findings? | §3.3; §3.4; Table 3 |
| 31 | Clarity of major themes | Were major themes clearly presented in the findings? | §3.3 (eight themes presented as subheadings, U-T1–U-T4 and P-T1–P-T4) |
| 32 | Clarity of minor themes | Is there a description of diverse cases or discussion of minor themes? | §3.3; §3.4; Supplementary File S7 |

† Item 22 (data saturation). Saturation was not used as a stopping criterion and is not claimed. Qualitative sample sufficiency was instead justified by information power (§2.3), considering the focused research aim, the specificity of two information-rich cohorts, and a structured, theory-informed analysis.

*Reference: Tong A, Sainsbury P, Craig J. Consolidated criteria for reporting qualitative research (COREQ): a 32-item checklist for interviews and focus groups. Int J Qual Health Care 2007;19(6):349–57. doi:10.1093/intqhc/mzm042*
