## Appendix B. Mixed Methods Appraisal Tool (MMAT), version 2018 for "Boora, an AI-assisted digital platform for overweight and obesity care in Brazilian primary care: a formative mixed-methods evaluation of perceived usability and acceptability" (from another version of this preprint: version 1, 2026-07-16)

| Methodological quality criteria | Yes | No | Can't tell | Comments (reported in section) |
|---|---|---|---|---|
| ***Screening questions (for all types)*** | | | | |
| S1. Are there clear research questions? | ✓ | | | The aim is clearly stated (§1; §2.1): to evaluate perceived usability, acceptability, and user experience of the Boora prototype, explicitly not clinical effectiveness. |
| S2. Do the collected data allow to address the research questions? | ✓ | | | SUS scores and semi-structured interviews with both cohorts address perceived usability, acceptability, and experience (§2.4; §2.5). |
| ***Category 1. Qualitative*** | | | | |
| 1.1. Is the qualitative approach appropriate to answer the research question? | ✓ | | | Codebook thematic analysis is appropriate for exploring perceptions of usability, acceptability, and anticipated workflow fit (§2.1; §2.5). |
| 1.2. Are the qualitative data collection methods adequate to address the research question? | ✓ | | | Semi-structured interviews after hands-on use, following guides covering usability, usefulness, barriers, facilitators, anticipated clinical utility, and integration into care routines (§2.4; Supplementary Files S5 and S6). |
| 1.3. Are the findings adequately derived from the data? | ✓ | | | Themes derived through a documented codebook process with an audit trail and independent secondary coding of a maximum-variation subsample (§2.5; Supplementary File S7). |
| 1.4. Is the interpretation of results sufficiently substantiated by data? | ✓ | | | Each theme is supported by identified verbatim quotations (§3.3). |
| 1.5. Is there coherence between qualitative data sources, collection, analysis and interpretation? | ✓ | | | A coherent chain runs from interview domains to codes, categories, themes, and integrated interpretation (§2.5; §3.3; §3.4). |

| Methodological quality criteria | Yes | No | Can't tell | Comments (reported in section) |
|---|---|---|---|---|
| ***Category 4. Quantitative descriptive*** | | | | |
| 4.1. Is the sampling strategy relevant to address the research question? | ✓ | | | Purposive formative sampling is relevant to prototype evaluation; sample size was defined pragmatically, with qualitative sufficiency guided by information power (§2.3). |
| 4.2. Is the sample representative of the target population? | | ✓ | | The purposive, single-municipality sample requiring smartphone ownership and internet access was not designed to be representative of the target primary care population. This selection limit and its effect on equity inferences are explicitly acknowledged (§2.3; §4, Limitations). |
| 4.3. Are the measurements appropriate? | ✓ | | | The SUS is a validated instrument, applied in a cross-culturally adapted Brazilian Portuguese version, with standard scoring (§2.4; §2.5; Supplementary File S8). |
| 4.4. Is the risk of nonresponse bias low? | | | ✓ | All 15 users and all 8 professionals completed the SUS with no missing items (§3.2). However, 3 of 11 professionals contacted did not respond (§2.3; Supplementary File S3), so nonresponse bias cannot be excluded. |
| 4.5. Is the statistical analysis appropriate to answer the research question? | ✓ | | | Analysis is descriptive (mean, SD, median, IQR, range) with no inferential between-group testing, a conservative choice appropriate to the small formative sample and to the differing exposure conditions (§2.5). |
| ***Category 5. Mixed methods*** | | | | |
| 5.1. Is there an adequate rationale for using a mixed methods design to address the research question? | ✓ | | | A socio-technical evaluation requires both standardised usability measurement and |

| Methodological quality criteria | Yes | No | Can’t tell | Comments (reported in section) |
|---|---|---|---|---|
| | | | | contextual understanding of anticipated workflow fit, justifying a convergent design (§2.1). |
| 5.2. Are the different components of the study effectively integrated to answer the research question? | ✓ | | | The strands were integrated through a joint display (§2.5; §3.4; Table 3). |
| 5.3. Are the outputs of the integration of qualitative and quantitative components adequately interpreted? | ✓ | | | Meta-inferences are typed and hedged; individual-level linkage was not performed and no causal or implementation effect is implied (§3.4; Table 3). |
| 5.4. Are divergences and inconsistencies between quantitative and qualitative results adequately addressed? | ✓ | | | Convergences, complementarities, and divergences are examined explicitly in the joint display, including favourable global usability masking a weak AI component (§3.4; Table 3). |
| 5.5. Do the different components of the study adhere to the quality criteria of each tradition of the methods involved? | | | ✓ | The qualitative component follows COREQ and codebook thematic analysis, and the quantitative component uses a validated instrument with appropriate descriptive analysis. Overall adherence is nonetheless bounded by the non-representative descriptive sample (criterion 4.2) and by the absence of objective usability-performance data (§4, Limitations), so no definitive judgement is claimed. |

*Reference: Hong QN, Pluye P, Fàbregues S, Bartlett G, Boardman F, Cargo M, et al. Mixed Methods Appraisal Tool (MMAT), version 2018. Montreal: McGill University; 2018. Registration of Copyright (#1148552), Canadian Intellectual Property Office, Industry Canada.*
