## Supplementary File S1. Technical specifications, development assistance, data governance, and data flows of the Boora prototype for "Boora, an AI-assisted digital platform for overweight and obesity care in Brazilian primary care: a formative mixed-methods evaluation of perceived usability and acceptability" (from another version of this preprint: version 1, 2026-07-16)

### 1. Evaluated build and development process

Boora was evaluated as a formative functional prototype during January and February 2026. The production deployment was first made available on 10 January 2026 at boora-app.vercel.app. The updated build used during the evaluation was the deployment available on 30 January 2026 at the same address. The prototype was not a certified medical device and was not integrated with the national electronic health record.

The platform was developed across successive versions through manual programming by FFSC. OpenAI Codex was used selectively for interface scaffolding, CSS and component refinement, code suggestions, and troubleshooting. The exact underlying Codex model snapshot was not recorded. FFSC reviewed, edited, executed, and tested the suggested code and retained responsibility for the system architecture, data model, core logic, security rules, clinical scope, and final implementation.

No participant records, interview transcripts, qualitative codes, SUS datasets, or identifiable research data were intentionally submitted to Codex. Codex was not used to recruit participants, determine eligibility, calculate study results, code interviews, generate themes, or make clinical decisions. This disclosure concerns use during prototype development; responsibility for all software behavior and research reporting remained with the authors.

### 2. Software stack and deployment

| Component | Technology/version | Role in the evaluated prototype |
|---|---|---|
| Patient-facing interface | JavaScript; React 19.2.3; Vite 8.0.0; Tailwind CSS 4.2.1 | Responsive progressive web application (PWA) accessed through the browser; no app-store installation required. |
| Professional dashboard | JavaScript; React 19.2.3; Vite 8.0.0; Tailwind CSS 4.2.1 | Web dashboard using synthetic data during professional usability tasks. |
| Client data library | Firebase JavaScript SDK 13.8.0, including Cloud Firestore client modules | Read/write operations were governed by authentication state and Firestore security rules. |
| Authentication | Firebase Authentication using email and password credentials | Firebase UID was used as the account identifier. Password handling was delegated to Firebase Authentication; plaintext passwords were not stored in the application database. |

| Component | Technology/version | Role in the evaluated prototype |
|---|---|---|
| Operational database | Google Cloud Firestore (NoSQL) | Primary persistent application and study-linked platform data repository. |
| Server-side logic | Firebase Cloud Functions and restricted server-side endpoints | Used for privileged operations and logic that should not be exposed in the browser. |
| Web hosting | Vercel | Hosted the web interface. Vercel was not the primary persistent data repository. Edge or deployment routing was not restricted to a Brazil-only region in the prototype. |
| Patient conversational model | Google Gemini 2.0 Flash-Lite through the Google Generative Language API | No local model, fine-tuning, project-specific training, retrieval-augmented generation, or web search. |
| Food search support | Open Food Facts public product database | Used for food/product lookup. Search terms, rather than participant identity fields, were intended to be transmitted. |
| Transactional email | Resend API; package version not recovered from the evaluated checkpoint | Server-side delivery of account-creation and invitation emails. Credentials and sender identity were supplied through RESEND_API_KEY and RESEND_FROM environment variables rather than the browser client. |
| Push notifications | Firebase Cloud Messaging through the Firebase Messaging web client; service worker; Web Push VAPID key | Optional browser/PWA notifications. The client checked messaging support, requested notification permission, registered /firebase-messaging-sw.js, obtained a Firebase Cloud Messaging (FCM) registration token with a VAPID public key, and associated the token with the authenticated patient. |

#### 2.1 Version and reproducibility notes

- The listed versions describe the evaluated January 2026 build and should not be interpreted as current versions of the respective packages.
- The source repository and dependency lockfile were not deposited in a public repository as part of this formative study.
- Objective task-performance telemetry and detailed application-use logs were not collected for the study.

### 3. Data categories and whether collection was required

| Data category | Examples | Required or optional |
|---|---|---|
| Authentication and access control | Email address, Firebase UID, authentication metadata, and assigned role | Required for account creation and access. Credential processing was managed by Firebase Authentication. |
| Pseudonymous identity | Study or platform code and account-to-role linkage | Required. Participant names were not used as the primary record identifier in the research database. |
| Core anthropometric information | Weight, height, body mass index, and dates of entries | Required only to the extent needed for core weight-monitoring functions and study eligibility/characterisation. Eligibility was verified separately in the electronic record. |
| Sociodemographic and professional characteristics | Age, sex, schooling, income category, profession, training, and experience variables | Collected for research characterisation. Not all questionnaire variables were required for routine platform operation. |
| Self-monitoring records | Dietary entries, water intake, physical activity, routine records, and weight history | Optional user-entered data. |
| Goals and care-support content | Goals, reminders, meal-plan links/documents, educational material access, and free-text notes | Optional, depending on the function used. |
| Conversational interactions | Messages entered into the patient assistant and the assistant responses | Optional. Only participants who chose to use the assistant generated these data. |

| Data category | Examples | Required or optional |
|---|---|---|
| Professional dashboard data | Synthetic patient profiles, risk flags, and task scenarios | Synthetic during the professional evaluation; professionals did not access records from the user cohort. |
| Interview and usability data | Audio recordings, transcripts, questionnaires, SUS item responses, and coded excerpts | Required for participants who enrolled in the research procedures, under informed consent; stored separately from routine authentication records where practicable. |
| Messaging and notification routing | Thread and message identifiers, sender role, timestamps, recipient email for transactional messages, notification-permission state, and FCM registration token/device metadata when push was enabled | Messaging records were generated when secure messaging was used. Email address was required for account access and transactional email. Push permission and FCM token registration were optional; declining notifications did not prevent platform use. |

**4. Storage, residence, access, and security controls**

| Control | Implementation in the evaluated prototype |
|---|---|
| Primary data residence | The Firebase project used for research data was configured in southamerica-east1 (Sao Paulo, Brazil). |
| Vercel hosting | Vercel hosted the web interface and was not the primary persistent repository. The prototype did not document or enforce a Brazil-only Vercel execution region. |
| Encryption in transit | Connections to Firebase, Vercel, and external APIs used provider-managed HTTPS/TLS. |
| Encryption at rest | Firestore records were protected through provider-managed encryption at rest. |
| Authentication | Centralised Firebase Authentication using email/password credentials and Firebase UIDs. |
| Authorisation | Role-based access control and Firestore security rules restricted data visibility and operations according to user role. |

| Control | Implementation in the evaluated prototype |
|---|---|
| Pseudonymisation | Application and research records were linked primarily to alphanumeric or Firebase identifiers rather than participant names. The code-to-identity key was restricted to authorised personnel. |
| Administrative access | Direct project-console or database access was restricted to authorised research and technical personnel. |
| Secrets | External API credentials were stored as server-side environment variables or provider-managed secrets and were not intentionally embedded in the client interface. |
| Auditability | The prototype did not implement a comprehensive clinical audit trail covering every read, write, export, and administrative action. |
| Independent assurance | No independent penetration test, privacy impact assessment, security certification, or external LGPD compliance audit was conducted. |
| Resend processing | Transactional email required transmission of the recipient email address, sender identity, subject, and message body to Resend. The prototype did not document or enforce Brazil-only processing or storage for this provider. |
| FCM/Web Push routing | Push delivery used an FCM registration token and a browser service worker. The VAPID public key was exposed to the client as intended for Web Push, whereas server credentials remained restricted. FCM routing and delivery were not confined to the Firestore southamerica-east1 data region. |
| Notification-content minimisation | Push and email alerts were secondary delivery channels rather than the source of truth; the message remained in the authenticated Firestore thread. Because notification previews may appear on lock screens or email clients, only generic notification text should be used and sensitive clinical content should remain inside the authenticated application. |

#### 5. Messaging, transactional email, and push notifications

The prototype included authenticated, Firestore-based professional-to-patient messaging and two ancillary notification mechanisms: transactional email through Resend and optional browser/PWA push notifications through FCM. The recovered implementation evidence was located in api/createProfessionalAccount.js, api/createPatientAccount.js, src/lib/push.js, the firebase-messaging service worker, and the Firestore-triggered notifyPatientOnProMessage function.

Resend was therefore part of the application architecture, not a development-only tool. FCM registration depended on browser support and explicit user permission. A newly created professional message in threads/{threadId}/messages/{msgId} activated a server-side notification workflow; push delivery depended on a current token and service-worker support, while email delivery depended on the configured Resend credentials and a valid recipient address.

| Element | Recovered implementation and interpretation |
|---|---|
| Source of truth | Messages were stored in authenticated Firestore threads. Email and push were notification channels and did not replace the in-application message record. |
| Transactional email | The Resend SDK/API was invoked server-side for account creation and invitation flows. Environment variables supplied the API key and verified sender. The Resend package version was not recoverable from the supplied checkpoint. |
| Push opt-in | The PWA checked whether Firebase Messaging was supported, requested browser notification permission, registered /firebase-messaging-sw.js, and used a VAPID public key to obtain an FCM token. Permission and token creation were optional. |
| Token storage and linkage | The FCM token function was tied to the authenticated Firebase user/patient identifier so that a notification could be routed to the correct browser or device. Tokens are routing identifiers and may become invalid after browser, permission, or device changes. |
| Message-triggered notification | A Firestore onDocumentCreated trigger monitored new documents under threads/{threadId}/messages/{msgId}. The recovered function was named notifyPatientOnProMessage and was intended to notify the patient when a professional created a new message. |
| Third-party data disclosure | Resend necessarily received the recipient email address and email payload. FCM necessarily received the registration token and notification-routing payload. These provider flows were outside the Firestore Brazil-region boundary. |
| Clinical monitoring | Notification generation did not constitute real-time clinical monitoring. During the 24-hour study exposure, entries and messages were not |

| Element | Recovered implementation and interpretation |
| --- | --- |
| | continuously reviewed for clinical escalation, and no clinical feedback was automatically generated. |
| Evaluation scope | The study did not measure delivery success, latency, bounce rates, token expiry, duplicate notifications, opt-out persistence, notification-preview confidentiality, or the effect of notifications on engagement. |

#### 6. Conversational AI data flows and safeguards

The patient assistant was an educational feature rather than a clinical decision system. A fixed Brazilian Portuguese instruction was prepended to the recent conversation context, limiting responses to general education on healthy habits and weight management, prohibiting diagnosis and medication advice, and directing warning signs to in-person or urgent care. The full instruction is reported in Supplementary File S2.

| Element | Data flow or safeguard |
| --- | --- |
| Input supplied by the user | Free-text message entered voluntarily in the assistant. |
| Context supplied to the model | The fixed guardrail instruction plus up to the 12 most recent messages needed to maintain conversational continuity. |
| Clinical database access | None. The patient assistant endpoint did not retrieve Firestore clinical records, anthropometric history, meal logs, or professional notes. |
| Retrieval and external search | No retrieval-augmented generation and no web search. |
| Actions | No function for writing clinical records, generating prescriptions, changing care plans, sending clinical alerts, or initiating clinical actions. |
| Personal identifiers | Users were instructed not to enter identifying information. The patient-facing endpoint did not include automated detection or guaranteed refusal/redaction of all identifiers. |
| Third-party processing | Message content sent to the Google Generative Language API could be processed outside the Firebase southamerica-east1 data region. The prototype did not configure a Brazil-only processing location for the external model API. |

| Element | Data flow or safeguard |
|---|---|
| Professional-facing assistant | The assistant shown during professional usability tasks was simulated. A separate authenticated backend existed for restricted, pseudonymous snapshots and selected-identifier redaction, but its model accuracy and clinical safety were not evaluated in this study. |
| Evaluation scope | The study assessed perceptions of usability, acceptability, and role clarity. It did not test systematic model accuracy, guardrail adherence, privacy leakage, or clinical safety. |

**7. Retention, deletion, and study monitoring**

- Study-linked platform records, research questionnaires, SUS data, audio recordings, transcripts, coding files, and the code-to-identity key are subject to the five-year retention period established in the approved protocol.
- Operational notification-routing data, including FCM registration tokens and delivery metadata, were not clinical outcomes. Tokens could be replaced or invalidated when permission, browser, or device state changed; the prototype did not implement a separately validated token-lifecycle or delivery-log retention policy.
- After the five-year period, the research team will securely delete the retained electronic research records and destroy the re-identification key, subject to applicable institutional and legal requirements.
- Deletion was a controlled research-administration procedure; the evaluated prototype did not provide participants with an automated self-service account erasure workflow.
- During the 24-hour user exposure, platform entries were not monitored clinically in real time. No individual clinical feedback was delivered through the platform, and entries were reviewed only after the exposure period to characterise use and support the interview.
- No platform-related clinical or safety event was reported during the study.
- 

**8. Interpretation and limitations of the technical description**

These controls describe a research prototype and its recovered architecture. They do not establish certified compliance with the Brazilian General Data Protection Law, conformity assessment, production-grade cybersecurity, or assured notification delivery. The study did not independently test Resend or FCM processing locations, email deliverability, service-worker behaviour across browsers, token revocation, lock-screen privacy, rate limiting, abuse prevention, message-notification duplication, or end-to-end delivery logs. The listed controls and provider flows should therefore be interpreted as implementation details and limitations of the evaluated prototype rather than as evidence of production readiness.
