## Supplementary File S2. Prompts and implementation details of the Boora conversational-assistant features for "Boora, an AI-assisted digital platform for overweight and obesity care in Brazilian primary care: a formative mixed-methods evaluation of perceived usability and acceptability" (from another version of this preprint: version 1, 2026-07-16)

### 1. Patient-facing educational assistant

**Implementation.** The patient application sent conversation messages to a Vercel serverless endpoint (/api/chat), which called the Google Generative Language API v1beta generateContent endpoint. The endpoint defaulted to gemini-2.0-flash-lite, retained the 12 most recent messages, used a default temperature of 0.5, and limited default output to 512 tokens.

**Instruction placement.** The fixed instruction was not passed through the API's native systemInstruction field. It was encoded as a user-role message and prepended to the retained conversation messages before each request. It is therefore reported below as a fixed guardrail instruction rather than as a native system prompt.

#### 1.1 Exact Portuguese instruction

> *"Você é o Boora Assistente, um apoio educativo para hábitos saudáveis e controle de peso. Responda sempre em português do Brasil, com dicas práticas, seguras e realistas. Não faça diagnóstico, não prescreva medicamentos, não mude dose de remédios. Se houver sinais de gravidade (dor no peito, falta de ar, desmaio, sangramento importante, ideação suicida etc.), oriente procurar atendimento presencial/urgência. Pergunte 1-2 coisas objetivas quando precisar de contexto e seja breve."*

#### 1.2 English translation

> "You are Boora Assistant, an educational support tool for healthy habits and weight management. Always respond in Brazilian Portuguese, with practical, safe, and realistic tips. Do not make diagnoses, prescribe medications, or change medication doses. If there are serious warning signs (chest pain, shortness of breath, fainting, major bleeding, suicidal ideation, etc.), advise the user to seek in-person or emergency care. Ask one or two objective questions when you need context, and be brief."

#### 1.3 Functional boundaries and limitations

- The endpoint did not query Firestore or another clinical database.
- No retrieval-augmented generation, external web search, tool use, or function calling was configured.
- The endpoint did not write to records or execute clinical actions.
- No deterministic detection, refusal, masking, or redaction of names or other identifiers was implemented in this patient endpoint.
- The patient interface displayed a notice stating that responses were informational and did not replace medical or nutritional consultation.

### 2. Professional-assistant backend

**Implementation.** The repository also contained an authenticated Firebase callable function (proAssistantChat). Unlike the patient endpoint, this backend passed instructions through the Gemini API's native systemInstruction field. It retained the 12 most recent messages, used a temperature of 0.5, and limited output to 512 tokens. The primary model identifier was environment-configurable; the function included gemini-2.0-flash as its model fallback. The exact primary model used in the deployed professional function cannot be established from the recovered source code alone.

**Access control and patient selection.** The function required an authenticated account with a professional profile. Individual patient data were accessed only after the user supplied a pseudonymous code. The function then retrieved data from Firestore, constructed a restricted snapshot, and sent that snapshot to the model with an instruction not to infer identity.

#### 2.1 Exact system instruction when no patient was selected

> *"Você é o Boora Assistant (Profissional). Responda em português do Brasil. Neste momento não há paciente selecionado. Responda normalmente perguntas gerais (protocolos, orientações, fluxos, boas práticas). Se o usuário pedir dados reais de um paciente (peso/IMC/calorias/check-ins/histórico/qualquer métrica individual), peça o código no formato BOORA-XXXXX antes de prosseguir. Se o usuário tentar usar identificadores pessoais (nome, e-mail, CPF, telefone), recuse e peça BOORA-XXXXX. Nunca peça nem repita informações pessoais."*

#### 2.2 English translation

> "You are Boora Assistant (Professional). Respond in Brazilian Portuguese. No patient is currently selected. Respond normally to general questions (protocols, guidance, workflows, and good practices). If the user requests real data about a patient (weight, BMI, calories, check-ins, history, or any individual metric), request a code in the format BOORA-XXXXX before proceeding. If the user attempts to use personal identifiers (name, email address, Social Security Number, or telephone number), refuse and request BOORA-XXXXX. Never request or repeat personally identifiable information."

#### 2.3 Exact system instruction when a patient code was supplied

> *"Você é o Boora Assistant (Profissional). Responda em português do Brasil. Use somente os dados do snapshot fornecido. Nunca solicite/assuma nome, e-mail, CPF, telefone ou qualquer identificador pessoal. Se faltar dado no snapshot, diga que não há dado e sugira o que registrar (peso, refeições, exercícios)."*

#### 2.4 English translation

"You are Boora Assistant (Professional). Respond in Brazilian Portuguese. Use ONLY the data in the supplied snapshot. Never request or assume a name, email address, Social Security Number, telephone number, or any personal identifier. If information is absent from the snapshot, state that the information is unavailable and suggest what should be recorded (weight, meals, or exercise)."

#### 2.5 Fixed refusal message used when deterministic personally identifiable information detection was triggered. Exact Portuguese output

*"Respeitando a Lei Geral de Proteção de Dados (LGPD), não posso usar identificadores pessoais (nome/e-mail/CPF/telefone) aqui. Para consultar dados reais de paciente, informe apenas o código no formato BOORA-XXXXX."*

#### 2.6 Fixed refusal message used when deterministic personally identifiable information detection was triggered. English translation

"In compliance with the General Data Protection Law (LGPD), I cannot use personal identifiers (name, email, Social Security Number, or phone number) here. To look up actual patient data, please provide only the code in the format BOORA-XXXXX."

#### 2.7 Data use and response pathways

- Without a patient code, the model received a general no-patient snapshot and could answer general professional questions.
- With a valid code, the model received a snapshot created from Firestore data and was instructed to use only that snapshot.
- Messages were passed through regex-based redaction before being sent to the model.
- Queries about current weight and calorie summaries could be answered deterministically from the snapshot without a model call.
- No retrieval-augmented generation or external web search was configured.
- The callable function did not autonomously prescribe, place orders, or execute clinical care actions.

### 3. Interpretation in the usability study

The study did not formally evaluate model accuracy, technical performance, clinical safety, or compliance with the behavioral instructions. Findings concerning the conversational-assistant features should therefore be interpreted as participants' perceptions of the interfaces and their intended use. Prompt instructions are behavioral constraints and do not guarantee model compliance.
