## Supplementary File S3. Recruitment and completion flow for the user and professional cohorts for "Boora, an AI-assisted digital platform for overweight and obesity care in Brazilian primary care: a formative mixed-methods evaluation of perceived usability and acceptability" (from another version of this preprint: version 1, 2026-07-16)

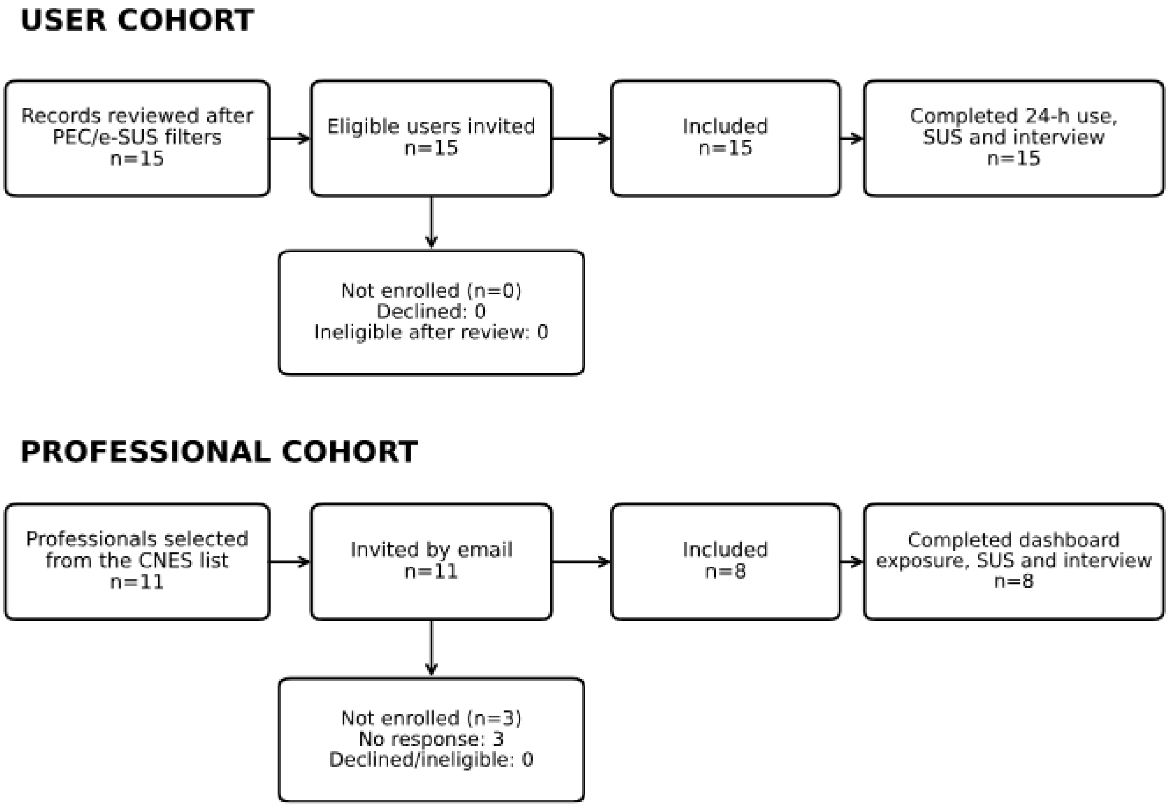


**Notes.** For the user cohort, the flow begins after PEC/e-SUS database filtering for a body mass index of 25 kg/m² or higher and exclusion of pregnancy. All 15 eligible users invited were included and completed the 24-hour application use period, System Usability Scale, and interview. Eleven professionals were invited sequentially from the National Registry of Health Establishments list; three did not respond within seven working days, and eight were included and completed the dashboard exposure, System Usability Scale, and interview.
