## Supplementary File S4. Standardised professional-dashboard exposure tasks for "Boora, an AI-assisted digital platform for overweight and obesity care in Brazilian primary care: a formative mixed-methods evaluation of perceived usability and acceptability" (from another version of this preprint: version 1, 2026-07-16)

**Purpose.** This file documents the structured exposure used to familiarise health professionals with the Boora dashboard before completion of the System Usability Scale and the subsequent semi-structured interview.

**Standardised exposure to the professional dashboard**

**Setting and duration.** Professionals accessed the dashboard by secure videoconference and were given approximately 20 minutes to explore predefined monitoring functions using exclusively synthetic patient data. No professional accessed information generated by the participating user cohort.

**Introductory instruction to participants.** "The data displayed in this prototype are synthetic and do not correspond to real patients. Please use the dashboard to complete the monitoring tasks below as you would when reviewing patients in routine primary care. There are no right or wrong clinical answers; the purpose is to allow you to experience the interface before answering the usability questionnaire and participating in the interview."

**Predefined monitoring tasks**

| Task | Action | Expected interface element |
|---|---|---|
| 1. Review the overview screen | Locate the summary indicators for active patients, goals achieved, dropout alerts, and goal-risk alerts. | Overview indicators |
| 2. Identify a patient requiring follow-up | Use the priority intervention queue to identify one synthetic patient flagged for closer monitoring. | Priority intervention queue |
| 3. Review the selected patient's anthropometric information | Open the patient record and locate the current weight, body mass index, and recent weight change. | Patient profile and weight/BMI fields |
| 4. Review recent self-monitoring information | Locate the available records on goals, food intake, physical activity, or check-ins for the selected patient. | Goals, dietary records, activity records, or check-ins |
| 5. Interpret the displayed risk flags | Identify whether dropout-risk or goal-risk flags are present and review the rule-based reason displayed for each flag. | Rule-based dropout- and goal-risk flags |
| 6. Prioritise a monitoring response | Based on the information shown, indicate which patient would be prioritised for follow-up and what type of follow-up would be considered in routine primary care. | Synthetic patient monitoring information |

| | | |
|---|---|---|
| 7. Inspect the communication and assistant area | Locate the messaging or educational-assistant area, review the privacy notice, and avoid entering any real patient identifier. | Messages/assistant panel and privacy notice |
| 8. Return to the overview | Identify any information or functionality that appeared missing, unnecessary, or potentially useful for longitudinal primary care follow-up. | Dashboard overview and navigation |

**Performance-data statement**

The task sequence was used only to structure exposure to the prototype. Task-completion times, completion rates, navigation errors, requests for assistance, and other objective usability-performance metrics were not recorded. The study evaluated perceived usability and participants' qualitative accounts.
