## Supplementary File S5. Semi-structured interview guide for professionals for "Boora, an AI-assisted digital platform for overweight and obesity care in Brazilian primary care: a formative mixed-methods evaluation of perceived usability and acceptability" (from another version of this preprint: version 1, 2026-07-16)

### Interviewer introduction

"Hello, and thank you again for your time and collaboration. This interview aims to obtain your professional perspective on the prototype of the Boora monitoring dashboard. Your critical assessment is fundamental to this project. As agreed in the informed-consent process, I will now begin the audio recording."

### Block 1. Overall impressions and interface

- What was your overall impression when interacting with the Boora professional dashboard?
- How did you perceive the presentation of information, including graphs, lists, and patient data?
- To what extent did the interface appear appropriate for clinical use?
- Probe: Could you describe in detail which aspects led you to this assessment?

### Block 2. Usability and workflow

- From a navigation perspective, what appeared most intuitive or easiest to locate on the dashboard?
- Did any functionality or information appear confusing, unnecessary, or difficult to access?
- Considering routine primary care practice, how could this dashboard be used? Would it save time or create additional workload?
- Probe: Could you describe this potential effect on your work routine in greater detail?

### Block 3. Clinical usefulness and relevance

- How clinically relevant were the user information displayed, such as dietary records and weight progression, to your decision-making process?
- How could data from this platform improve therapeutic planning and longitudinal follow-up for users with overweight or obesity?
- How would you feel about using data generated by this platform as a complement to information obtained during consultations?
- How do you view a hypothetical integration of Boora data with the PEC/e-SUS APS electronic health record?
- Probe: Could you describe the potential effect of this integration on your work routine in greater detail?

### Block 4. Suggestions and closing questions

- If you could redesign or adapt the dashboard for your daily practice, what would be the main changes you would make?
- In your view, what other functions or types of data, such as reports or alerts, could make this tool more valuable to the healthcare team?
- Do you have any other comments, criticisms, or suggestions? Is there anything relevant that we have not discussed?

### Closing script

"Thank you very much for your detailed assessment. Your professional contribution is essential to improving this tool. I am now ending the recording. Thank you."
