## Supplementary File S6. Semi-structured interview guide for user participants for "Boora, an AI-assisted digital platform for overweight and obesity care in Brazilian primary care: a formative mixed-methods evaluation of perceived usability and acceptability" (from another version of this preprint: version 1, 2026-07-16)

**Interviewer introduction**

"Hello, and thank you again for taking part. As we discussed, this interview is the final stage of our study. Its purpose is to hear your views about your experience of using the Boora application over the previous 24 hours. There are no right or wrong answers, and this conversation will remain confidential. As authorised in the informed consent form, I am now starting the audio recording."

**Block 1. Overall impressions and initial use**

- To begin, how has your day been so far?
- How would you describe your overall experience of using the Boora application since yesterday?
- What did you think about the installation process and the initial instructions you received? What were the first few hours of using the application like?
- What was your first impression of the application's appearance and design?

**Block 2. Usability and ease of use in everyday life**

- Thinking about the activities you completed, such as recording your meals or mood, what was easiest to do?
- Did you experience any difficulties? Was there any point at which you felt confused or the application did not work as expected?
- ***Probe:*** *Could you explain in more detail what happened at that moment?*
- What was it like to interact with the virtual assistant (chatbot)? Were its responses clear and useful to you?
- ***Probe:*** *What had you expected the responses to be like?*

**Block 3. Usefulness and personal relevance**

- How could this application actually help you take better care of your health in everyday life? In what ways?
- Of the features you explored, which do you think would be most useful to you in the long term? Which would be least useful?
- What would it be like for you to use this application every day?
- ***Probe:*** *What would motivate, or discourage, you from continuing to use a tool like this every day?*

**Block 4. Suggestions and closing questions**

- If you could change anything about the Boora application, what would it be?
- Was there any feature or information that you felt was missing and considered important?
- Would you like to add any other comments or offer any final suggestions?

**Closing script**

"Your views have been extremely valuable to our project. Thank you very much for your time and contribution. I am now ending the recording. Thank you."
