## Supplementary File S7. Consolidated codebook and code-category-theme hierarchy for "Boora, an AI-assisted digital platform for overweight and obesity care in Brazilian primary care: a formative mixed-methods evaluation of perceived usability and acceptability" (from another version of this preprint: version 1, 2026-07-16)

**Scope.** This consolidated codebook documents the final thematic hierarchy used for 15 user interviews (180 coded excerpts) and eight professional interviews (128 coded excerpts). Four final themes were generated for each cohort. Operational definitions and illustrative excerpts were translated from Brazilian Portuguese into English for publication.

**Consolidation rule.** The source codebooks contained broad descriptive categories, including cross-cutting categories for barriers and suggested improvements. For the final code → category → theme hierarchy, these categories were consolidated or partitioned according to the analytic meaning of each code. Code labels and operational boundaries were retained, while each substantive code was assigned once to a final category and theme. The provisional "emergent finding" codes were retained as analytic-management codes and were not treated as final themes.

**Interpretive use.** Illustrative excerpts clarify coding boundaries; they are not intended to represent frequency or prevalence. The coded-excerpt totals refer to the analytic corpora supplied by the authors; code-level frequency counts were not included in the source codebooks.

| Cohort | Interviews | Coded excerpts | Final themes | Codebook codes |
|---|---|---|---|---|
| Users | 15 | 180 | 4 | 35 (34 thematic + 1 provisional) |
| Professionals | 8 | 128 | 4 | 23 (22 thematic + 1 provisional) |

#### 1. Overview of final themes and consolidated categories

##### 1.1 User cohort

| Theme | Consolidated category | Operational definition |
|---|---|---|
| U-T1. Onboarding and routine logging burden<br>Captures first contact with the application and the effort required for everyday interaction, ranging from smooth onboarding and simple entries to burdensome detailed logging and interface friction. | **U1-A. Initial impressions and onboarding** | Initial visual acceptance, installation, instructions, and the participant's first overall appraisal of the application. |
| | **U1-B. Low-friction navigation and simple logs** | Ease of finding functions and completing brief, low-complexity self-monitoring entries. |
| | **U1-C. Burden and friction in detailed logging** | Cognitive, practical, content, and performance barriers encountered when recording food, portions, exercise, or multiple daily fields. |

| Theme | Consolidated category | Operational definition |
|---|---|---|
| | **U1-D. Interface and workflow refinements** | Suggestions to simplify interaction, standardise installation, or make data easier to export and share. |
| U-T2. A useful but limited AI assistant<br>Captures how users understood and valued the conversational assistant, including perceived clarity, limited use or responsiveness, and expectations for more tailored nutrition and activity content. | **U2-A. Perceived value and limitations of the assistant** | Positive experiences with the assistant as well as confusion, limited use, generic responses, or unmet expectations. |
| | **U2-B. Desired personalised content** | Requests for culturally relevant dietary guidance, recipes, and exercise content adapted to individual needs or limitations. |
| U-T3. Engagement through feedback, accountability, and self-management<br>Captures mechanisms that support or undermine continued use, including progress feedback, habit awareness, professional accountability, motivation, time pressure, and selective adherence. | **U3-A. Feedback and self-monitoring** | Use of weight, hydration, habits, and goals to make progress visible and support day-to-day self-care. |
| | **U3-B. Accountability and motivation** | Motivation arising from perceived improvement and awareness that health professionals may review the records. |
| | **U3-C. Threats to sustained use** | Fatigue, competing demands, limited time, and preferences for partial or selective use that may reduce continuity. |
| | **U3-D. Supports for sustained engagement** | Requested reminders and broader self-monitoring or device connections intended to sustain use and increase personal relevance. |
| U-T4. Digital inclusion as a condition for sustained use<br>Captures whether users could independently access and use the application given digital literacy, accessibility, connectivity, and device constraints. | **U4-A. Assisted onboarding and digital literacy** | Dependence on other people or guided learning resources to install, understand, or operate the application. |

| Theme | Consolidated category | Operational definition |
|---|---|---|
| | **U4-B. Accessible design and communication** | Need for readable text, clear controls, plain language, audio, and design suitable for older or less digitally experienced users. |
| | **U4-C. Connectivity and device constraints** | Limitations arising from mobile data, internet access, device performance, and the need for lightweight or offline operation. |

**1.2 Professional cohort**

| Theme | Consolidated category | Operational definition |
|---|---|---|
| P-T1. Interface fit for primary care practice<br>Captures initial acceptance, visual clarity, navigation, perceived clinical appropriateness, and the extent to which the dashboard appeared applicable to routine primary care. | **P1-A. Initial acceptance and interface clarity** | Overall impressions of the dashboard's appearance, organisation, navigation, and suitability for clinical use. |
| | **P1-B. Applicability and refinement** | Overall appraisal of real-world potential and the need for adjustments before broader adoption. |
| P-T2. Panel-managed care and longitudinal monitoring<br>Captures the dashboard's perceived value for stratifying risk, prioritising patients, managing a care panel, monitoring progress, and supporting professional decisions and actions. | **P2-A. Risk stratification and prioritisation** | Identification of priority patients, low adherence, dropout risk, and opportunities for proactive follow-up. |
| | **P2-B. Longitudinal and panel management** | Continuous follow-up, workflow efficiency, consolidation of information, and planned management of groups of patients. |
| | **P2-C. Decision support and professional action** | Use of guidelines and patient-generated information to improve clinical reasoning and professional decision-making. |
| | **P2-D. Requested clinical management functions** | Suggested functions for dietary plans, clinical actions, goals, individual care plans, and broader clinical information. |

| Theme | Consolidated category | Operational definition |
|---|---|---|
| P-T3. Conditions for safe implementation<br>Captures organisational, educational, and governance conditions considered necessary for responsible adoption without adding unmanageable workload or poorly defined AI-mediated communication. | **P3-A. Work organisation and implementation support** | Need for protected time, clear responsibilities, training, tutorials, and guided implementation. |
| | **P3-B. AI and chat governance** | Need to define the assistant's role, operational limits, safety rules, and appropriate use of messaging and patient data. |
| P-T4. Systemic integration as the basis for continuity of care<br>Captures interoperability with existing health-information systems and the role of shared, traceable records in reducing duplicate work and supporting multiprofessional continuity. | **P4-A. Interoperability and reduction of duplicate work** | Integration with PEC/e-SUS or other systems to avoid repeated entry, transcription, and fragmented screens. |
| | **P4-B. Multiprofessional continuity and coordination** | Shared information and aligned goals across professionals and encounters. |
| | **P4-C. Information governance and official record** | Security, traceability, and the legitimacy of formal records in shared care. |

### 2. Hierarchical tree: code → category → theme

#### 2.1 User cohort

**U-T1 → Onboarding and routine logging burden**
↳ U1-A → Initial impressions and onboarding
↳ U01 → Overall positive experience
↳ U02 → Clean, organised, and pleasant interface
↳ U03 → Smooth installation and clear instructions
↳ U1-B → Low-friction navigation and simple logs
↳ U05 → General ease of use and navigation
↳ U06 → Simple weight logging
↳ U07 → Simple water logging
↳ U1-C → Burden and friction in detailed logging
↳ U08 → Burdensome or confusing food logging
↳ U09 → Insufficient food database and regional foods
↳ U10 → Difficulty with measures and portions
↳ U11 → Confusing or impractical exercise logging
↳ U12 → Excess functions, fields, and completion burden
↳ U13 → Slowness, loading, or freezing
↳ U1-D → Interface and workflow refinements
↳ U29 → Simplification of screens and flows
↳ U34 → Standard installation, data sharing, or export

**U-T2 → A useful but limited AI assistant**
↳ U2-A → Perceived value and limitations of the assistant
↳ U25 → Assistant perceived as useful or clear
↳ U26 → Assistant underused, confusing, or insufficiently dynamic
↳ U2-B → Desired personalised content
↳ U27 → Dietary guidance, recipes, and regional foods
↳ U28 → Adapted exercises and suitability of videos

**U-T3 → Engagement through feedback, accountability, and self-management**
↳ U3-A → Feedback and self-monitoring
↳ U14 → Visual monitoring of weight and progress
↳ U15 → Hydration and habit monitoring
↳ U16 → Self-care, goals, and self-monitoring
↳ U3-B → Accountability and motivation
↳ U17 → Professional follow-up as a motivator
↳ U18 → Engagement and motivation to use
↳ U3-C → Threats to sustained use
↳ U19 → Demotivation and risk of discontinuation
↳ U20 → Selective use or partial adherence
↳ U21 → Lack of time and busy routines
↳ U3-D → Supports for sustained engagement
↳ U31 → Active notifications and reminders
↳ U33 → Recording clinical parameters and device integration

**U-T4 → Digital inclusion as a condition for sustained use**
↳ U4-A → Assisted onboarding and digital literacy

↳ U04 → Installation or initial use with third-party support
↳ U22 → Low digital familiarity and dependence on help
↳ U30 → Tutorial, step-by-step guidance, audio, or supported implementation
↳ U4-B → Accessible design and communication
↳ U23 → Visual accessibility, plain language, and digital inclusion
↳ U4-C → Connectivity and device constraints
↳ U24 → Limited internet, mobile data, or low-specification phone
↳ U32 → Lightweight or offline functionality

**Provisional residual code**

↳ U35 → Emergent finding (not assigned to a final theme)

#### 2.2 Professional cohort

**P-T1 → Interface fit for primary care practice**

↳ P1-A → Initial acceptance and interface clarity
↳ P01 → Overall positive experience
↳ P02 → Clarity, visual organisation, and objectivity
↳ P03 → Intuitive navigation
↳ P04 → Suitability for clinical use
↳ P1-B → Applicability and refinement
↳ P21 → Real-world potential in primary care
↳ P22 → General suggestions and platform refinements

**P-T2 → Panel-managed care and longitudinal monitoring**

↳ P2-A → Risk stratification and prioritisation
↳ P05 → Rapid identification of priority patients
↳ P09 → Active outreach, reminders, and prevention of dropout
↳ P2-B → Longitudinal and panel management
↳ P06 → Longitudinal monitoring
↳ P07 → Workflow optimisation
↳ P08 → Centralisation and management of the care panel
↳ P2-C → Decision support and professional action
↳ P10 → Protocols as decision support
↳ P11 → Improvement of professional decision-making
↳ P2-D → Requested clinical management functions
↳ P15 → Structured dietary prescription
↳ P16 → Recording clinical actions, goals, and individual care plans
↳ P17 → Expansion of patient clinical data

**P-T3 → Conditions for safe implementation**

↳ P3-A → Work organisation and implementation support
↳ P12 → Potential workload and protected time
↳ P13 → Training, tutorials, and system explanations
↳ P3-B → AI and chat governance
↳ P14 → Governance and scope of the assistant/chat

**P-T4 → Systemic integration as the basis for continuity of care**

↳ P4-A → Interoperability and reduction of duplicate work
↳ P18 → Interoperability with PEC/e-SUS and reduction of duplicate work
↳ P4-B → Multiprofessional continuity and coordination

↳ P19 → Continuity and multiprofessional communication

↳ P4-C → Information governance and official record

↳ P20 → Information security and official record

**Provisional residual code**

↳ P23 → Emergent finding (not assigned to a final theme)

### 3. Detailed user codebook

#### U-T1. Onboarding and routine logging burden

**Theme definition:** Captures first contact with the application and the effort required for everyday interaction, ranging from smooth onboarding and simple entries to burdensome detailed logging and interface friction.

#### U1-A. Initial impressions and onboarding

**Category definition:** Initial visual acceptance, installation, instructions, and the participant's first overall appraisal of the application.

| ID | Code | Operational definition | Illustrative excerpt (translated) |
|---|---|---|---|
| U01 | **Overall positive experience** | Apply when the participant reports a favourable overall impression, initial satisfaction, perceived usefulness, or good acceptance of the application. | *"Overall, I thought the application was very good; it has potential."* |
| U02 | **Clean, organised, and pleasant interface** | Apply when the participant describes the application as attractive, visually clean, well organised, easy to read, or having well-positioned buttons and information. | *"I thought it was attractive and organised."* |
| U03 | **Smooth installation and clear instructions** | Apply when the participant reports installing and starting the application without errors and understanding the initial instructions. | *"I downloaded and installed it quickly, and the instructions when the app opens are very clear."* |

#### U1-B. Low-friction navigation and simple logs

**Category definition:** Ease of finding functions and completing brief, low-complexity self-monitoring entries.

| ID | Code | Operational definition | Illustrative excerpt (translated) |
|---|---|---|---|
| U05 | **General ease of use and navigation** | Apply when the participant reports understanding the application, navigating it, locating functions, or using features without substantial difficulty. | *"It was a very good experience; nothing froze, and I was never unsure where to click."* |

| ID | Code | Operational definition | Illustrative excerpt (translated) |
|---|---|---|---|
| U06 | **Simple weight logging** | Apply when the participant describes weight entry as easy, quick, or requiring little effort. | *“Recording my weight was simple, no mystery.”* |
| U07 | **Simple water logging** | Apply when the participant describes water or hydration entry as easy, usually through a direct button or quick action. | *“The water part was very easy. You just press the little button.”* |

**U1-C. Burden and friction in detailed logging**

**Category definition:** Cognitive, practical, content, and performance barriers encountered when recording food, portions, exercise, or multiple daily fields.

| ID | Code | Operational definition | Illustrative excerpt (translated) |
|---|---|---|---|
| U08 | **Burdensome or confusing food logging** | Apply when the participant describes meal recording as tiring, slow, overly detailed, bureaucratic, confusing, or difficult to sustain in daily life. | *“When it came to food, I did not understand anything.”* |
| U09 | **Insufficient food database and regional foods** | Apply when the participant cannot find specific foods, reports failed food searches, or requests commonly consumed local or regional foods. | *“I would add more of our foods, regional things.”* |
| U10 | **Difficulty with measures and portions** | Apply when the participant is uncertain about quantities, grams, household measures, or how to estimate consumed portions. | *“I do not know how many grams there are in what I put on my plate.”* |
| U11 | **Confusing or impractical exercise logging** | Apply when the participant has difficulty adding exercises, understanding the activity-logging logic, or completing training details. | *“It took me a while to understand how to record the exercises.”* |

| ID | Code | Operational definition | Illustrative excerpt (translated) |
|---|---|---|---|
| U12 | **Excess functions, fields, and completion burden** | Apply when the participant reports too many screens, buttons, functions, fields, or information, resulting in confusion, fatigue, or difficulty maintaining daily completion. | *"There is too much to enter; I felt a bit dizzy with so many places to write things."* |
| U13 | **Slowness, loading, or freezing** | Apply when the participant reports slow loading, failed searches, phone freezing, or other performance limitations that interfere with use. | *"My phone is not one of the best; it takes a while to open and load things."* |

**U1-D. Interface and workflow refinements**

**Category definition:** Suggestions to simplify interaction, standardise installation, or make data easier to export and share.

| ID | Code | Operational definition | Illustrative excerpt (translated) |
|---|---|---|---|
| U29 | **Simplification of screens and flows** | Apply when the participant recommends reducing screens, clicks, fields, functions, or information to make the application simpler, faster, and more direct. | *"I would make the home screen simpler, with fewer things."* |
| U34 | **Standard installation, data sharing, or export** | Apply when the participant requests availability in an app store, a more conventional installation process, or options to download, export, or share graphs and data. | *"I would also like an option to download the graphs."* |

**U-T2. A useful but limited AI assistant**

**Theme definition:** Captures how users understood and valued the conversational assistant, including perceived clarity, limited use or responsiveness, and expectations for more tailored nutrition and activity content.

**U2-A. Perceived value and limitations of the assistant**

**Category definition:** Positive experiences with the assistant as well as confusion, limited use, generic responses, or unmet expectations.

| ID | Code | Operational definition | Illustrative excerpt (translated) |
|---|---|---|---|
| U25 | **Assistant perceived as useful or clear** | Apply when the participant reports that the chatbot responds clearly, provides guidance, answers questions, or offers rapid support. | *"The answers were clear; it responds properly."* |
| U26 | **Assistant underused, confusing, or insufficiently dynamic** | Apply when the participant did not use or understand the assistant, or considered it confusing, generic, shallow, unnecessary, or less useful than other functions. | *"I thought it would be more dynamic and help more."* |

**U2-B. Desired personalised content**

**Category definition:** Requests for culturally relevant dietary guidance, recipes, and exercise content adapted to individual needs or limitations.

| ID | Code | Operational definition | Illustrative excerpt (translated) |
|---|---|---|---|
| U27 | **Dietary guidance, recipes, and regional foods** | Apply when the participant requests dietary examples, guidance on what to eat, healthy recipes, ready-made meal suggestions, regional foods, or nutritional personalisation. | *"I would add more of our foods and some diet options for my profile."* |
| U28 | **Adapted exercises and suitability of videos** | Apply when the participant requests simpler exercises, adaptations for pain or functional limitations, or considers exercise videos difficult to access or unsuitable. | *"The least useful part was those training videos."* |

**U-T3. Engagement through feedback, accountability, and self-management**

**Theme definition:** Captures mechanisms that support or undermine continued use, including progress feedback, habit awareness, professional accountability, motivation, time pressure, and selective adherence.

**U3-A. Feedback and self-monitoring**

**Category definition:** Use of weight, hydration, habits, and goals to make progress visible and support day-to-day self-care.

| ID | Code | Operational definition | Illustrative excerpt (translated) |
|---|---|---|---|
| U14 | **Visual monitoring of weight and progress** | Apply when the participant values tracking weight over time, viewing graphs, noticing weight reduction, or seeing concrete progress. | *"The most useful part is definitely recording and following our weight progress, seeing the graph improve."* |
| U15 | **Hydration and habit monitoring** | Apply when the participant associates the application with remembering water intake, monitoring habits, or becoming aware of everyday health behaviours. | *"It helps a lot with awareness, especially of weight and water intake."* |
| U16 | **Self-care, goals, and self-monitoring** | Apply when the participant relates the application to improved self-care, focus on treatment, discipline, remembering goals, or monitoring their own behaviour. | *"Having this control on the phone helps us keep a closer watch on ourselves."* |

**U3-B. Accountability and motivation**

**Category definition:** Motivation arising from perceived improvement and awareness that health professionals may review the records.

| ID | Code | Operational definition | Illustrative excerpt (translated) |
|---|---|---|---|
| U17 | **Professional follow-up as a motivator** | Apply when the participant feels more motivated because a physician or health team may review their records or progress. | *"When we see the graph, we stop and think that we will have to show a result to the doctor."* |
| U18 | **Engagement and motivation to use** | Apply when the participant expresses willingness or intention to continue using the application, enthusiasm, perceived results, or other factors that increase adherence. | *"What encourages me is seeing that I am improving and managing to take care of myself."* |

**U3-C. Threats to sustained use**

**Category definition:** Fatigue, competing demands, limited time, and preferences for partial or selective use that may reduce continuity.

| ID | Code | Operational definition | Illustrative excerpt (translated) |
|---|---|---|---|
| U19 | **Demotivation and risk of discontinuation** | Apply when the participant describes factors that discourage continued use, cause fatigue, or increase the likelihood of abandoning the application. | *"What discourages me is having to fill in all this information every day; it gets tiring."* |
| U20 | **Selective use or partial adherence** | Apply when the participant would use only selected functions, use the application only on some days, require help, or favour only quick entries. | *"I would use it if my daughter were nearby to give me some guidance."* |
| U21 | **Lack of time and busy routines** | Apply when difficulty using the application is attributed to work, study, children, household duties, family responsibilities, or insufficient time. | *"Today has been very hectic, with work and many things to deal with."* |

**U3-D. Supports for sustained engagement**

**Category definition:** Requested reminders and broader self-monitoring or device connections intended to sustain use and increase personal relevance.

| ID | Code | Operational definition | Illustrative excerpt (translated) |
|---|---|---|---|
| U31 | **Active notifications and reminders** | Apply when the participant requests reminders for water, meals, weight, opening the application, or maintaining engagement. | *"I would use it every day, but I would need reminders, like phone notifications."* |
| U33 | **Recording clinical parameters and device integration** | Apply when the participant requests blood pressure, laboratory results, other health data, digital-scale integration, body composition, or connection to external devices. | *"I missed being able to record my blood pressure and test results."* |

**U-T4. Digital inclusion as a condition for sustained use**

**Theme definition:** Captures whether users could independently access and use the application given digital literacy, accessibility, connectivity, and device constraints.

**U4-A. Assisted onboarding and digital literacy**

**Category definition:** Dependence on other people or guided learning resources to install, understand, or operate the application.

| ID | Code | Operational definition | Illustrative excerpt (translated) |
|---|---|---|---|
| U04 | **Installation or initial use with third-party support** | Apply when the participant needed help from a family member, acquaintance, community health worker, or another person to install, start, or understand the application. | *"My daughter installed it; on my own, I would not even have known where to start."* |
| U22 | **Low digital familiarity and dependence on help** | Apply when the participant expresses digital insecurity, fear of making mistakes, difficulty handling technology, or dependence on help to install, navigate, or record information. | *"On my own, I would struggle a lot to download it."* |
| U30 | **Tutorial, step-by-step guidance, audio, or supported implementation** | Apply when the participant recommends a tutorial, explanatory video, audio guidance, an initial walkthrough, or in-person assistance to learn the application. | *"A video teaching how to use it very slowly would also be good."* |

**U4-B. Accessible design and communication**

**Category definition:** Need for readable text, clear controls, plain language, audio, and design suitable for older or less digitally experienced users.

| ID | Code | Operational definition | Illustrative excerpt (translated) |
|---|---|---|---|
| U23 | **Visual accessibility, plain language, and digital inclusion** | Apply when the participant requests larger text, improved legibility, clearer buttons, audio, simpler language, or adaptation for older adults or people with limited digital familiarity. | *"Make it easier for people like us who are older and do not understand these things very well."* |

**U4-C. Connectivity and device constraints**

**Category definition:** Limitations arising from mobile data, internet access, device performance, and the need for lightweight or offline operation.

| ID | Code | Operational definition | Illustrative excerpt (translated) |
|---|---|---|---|
| U24 | **Limited internet, mobile data, or low-specification phone** | Apply when the participant mentions limited mobile data, internet access, phone performance, freezing, or a need for lighter or offline operation. | *"I do not have spare internet data on my phone; watching a video uses it all up."* |
| U32 | **Lightweight or offline functionality** | Apply when the participant recommends operation without continuous internet, lower data use, faster loading, or compatibility with basic phones. | *"There should be a way to use it without needing internet all the time."* |

**4. Detailed professional codebook**

**P-T1. Interface fit for primary care practice**

**Theme definition:** Captures initial acceptance, visual clarity, navigation, perceived clinical appropriateness, and the extent to which the dashboard appeared applicable to routine primary care.

**P1-A. Initial acceptance and interface clarity**

**Category definition:** Overall impressions of the dashboard's appearance, organisation, navigation, and suitability for clinical use.

| ID | Code | Operational definition | Illustrative excerpt (translated) |
|---|---|---|---|
| P01 | **Overall positive experience** | Apply when the participant reports a favourable overall impression, initial satisfaction, good acceptance, or a positive overall evaluation of the professional dashboard. | *"The experience with the platform was good, and in the future it could be implemented in primary care routines."* |
| P02 | **Clarity, visual organisation, and objectivity** | Apply when the participant describes the interface as clean, organised, objective, clear, well distributed, uncluttered, or presenting information directly. | *"I found the platform objective, with good visual organisation."* |
| P03 | **Intuitive navigation** | Apply when the participant can locate modules, functions, screens, lists, patients, messages, or protocols without additional support. | *"I could easily access the patient, messaging, and protocol modules."* |
| P04 | **Suitability for clinical use** | Apply when the participant considers the dashboard compatible with clinical use, routine care, primary care, or follow-up of people with obesity. | *"The interface seems suitable for clinical use, mainly because it organises information within our daily workflow."* |

**P1-B. Applicability and refinement**

**Category definition:** Overall appraisal of real-world potential and the need for adjustments before broader adoption.

| ID | Code | Operational definition | Illustrative excerpt (translated) |
|---|---|---|---|
| P21 | **Real-world potential in primary care** | Apply when the participant considers the tool promising, applicable, relevant, or capable of concrete use in everyday primary care. | *"I consider the platform an excellent and functional idea, with potential to improve continuity of care in primary care."* |
| P22 | **General suggestions and platform refinements** | Apply when the participant recognises the platform's potential but identifies adjustments, refinements, improvements, or additions required before broader adoption. | *"I consider the platform relevant to primary care, with real potential for team use, although it still needs some adjustments."* |

**P-T2. Panel-managed care and longitudinal monitoring**

**Theme definition:** Captures the dashboard's perceived value for stratifying risk, prioritising patients, managing a care panel, monitoring progress, and supporting professional decisions and actions.

**P2-A. Risk stratification and prioritisation**

**Category definition:** Identification of priority patients, low adherence, dropout risk, and opportunities for proactive follow-up.

| ID | Code | Operational definition | Illustrative excerpt (translated) |
|---|---|---|---|
| P05 | **Rapid identification of priority patients** | Apply when the participant values alerts, intervention queues, risk status, unmet goals, or rapid identification of patients requiring priority attention. | *"I liked the rapid view of patients at greater risk. It makes everyday decision-making much easier."* |
| P09 | **Active outreach, reminders, and prevention of dropout** | Apply when the participant links the dashboard to identifying missed appointments, low adherence, dropout risk, reminders, returns, or proactive team contact. | *"The platform is useful for risk stratification, follow-up, and prioritising patients with a greater chance of dropout or deterioration."* |

**P2-B. Longitudinal and panel management**

**Category definition:** Continuous follow-up, workflow efficiency, consolidation of information, and planned management of groups of patients.

| ID | Code | Operational definition | Illustrative excerpt (translated) |
|---|---|---|---|
| P06 | **Longitudinal monitoring** | Apply when the participant values continuous follow-up of weight, BMI, adherence, clinical progress, or patients over time. | *"The platform is useful for care surveillance and following the patient over time."* |
| P07 | **Workflow optimisation** | Apply when the participant reports that the dashboard could improve the efficiency of consultations, follow-up, triage, prioritisation, or data organisation. | *"The concise presentation of key indicators and adherence makes it much easier to use in daily practice."* |
| P08 | **Centralisation and management of the care panel** | Apply when the participant refers to organising the care pathway, managing a patient panel, consolidating dispersed information, or planning follow-up for groups of patients. | *"Having this dashboard helps manage not only the individual but also the group."* |

**P2-C. Decision support and professional action**

**Category definition:** Use of guidelines and patient-generated information to improve clinical reasoning and professional decision-making.

| ID | Code | Operational definition | Illustrative excerpt (translated) |
|---|---|---|---|
| P10 | **Protocols as decision support** | Apply when the participant values protocols, guidelines, or reference materials as support for consultations, follow-up, or decision-making. | *"I navigated without difficulty through the patient list, the synthetic patient data, and the guidelines and protocols."* |
| P11 | **Improvement of professional decision-making** | Apply when the participant indicates that platform data could increase the safety, precision, or quality of clinical decisions, treatment adjustments, or professional actions. | *"The physician would have greater confidence when adjusting medications, for example, by knowing exactly what eating behaviour was recorded in the app."* |

**P2-D. Requested clinical management functions**

**Category definition:** Suggested functions for dietary plans, clinical actions, goals, individual care plans, and broader clinical information.

| ID | Code | Operational definition | Illustrative excerpt (translated) |
|---|---|---|---|
| P15 | **Structured dietary prescription** | Apply when the participant requests structured entry, editing, personalisation, or management of dietary plans within the platform. | *"I noticed a limitation in personalising the dietary prescription."* |
| P16 | **Recording clinical actions, goals, and individual care plans** | Apply when the participant recommends fields for clinical actions, guidance, therapeutic goals, individual care plans, agreed targets, or a history of interactions between the patient and team. | *"It would be useful to include a space to record clinical actions and link them to therapeutic goals."* |
| P17 | **Expansion of patient clinical data** | Apply when the participant requests laboratory tests, comorbidities, medications, blood pressure, glycated haemoglobin, clinical history, or other relevant patient parameters. | *"It would be relevant to integrate parameters such as blood pressure, glycated haemoglobin, and other comorbidities."* |

**P-T3. Conditions for safe implementation**

**Theme definition:** Captures organisational, educational, and governance conditions considered necessary for responsible adoption without adding unmanageable workload or poorly defined AI-mediated communication.

**P3-A. Work organisation and implementation support**

**Category definition:** Need for protected time, clear responsibilities, training, tutorials, and guided implementation.

| ID | Code | Operational definition | Illustrative excerpt (translated) |
|---|---|---|---|
| P12 | **Potential workload and protected time** | Apply when the participant identifies a risk of additional workload, excessive tasks, unclear responsibilities, or the need for protected time to use the dashboard. | *"The risk is that it becomes one more thing to open without protected time in the schedule."* |

| ID | Code | Operational definition | Illustrative excerpt (translated) |
|---|---|---|---|
| P13 | **Training, tutorials, and system explanations** | Apply when the participant mentions a learning curve, training needs, tutorials, guided implementation, more visible explanations, or clarification of functions and alerts. | *"It would be useful to include a brief tutorial within the system, especially to facilitate initial use."* |

**P3-B. AI and chat governance**

**Category definition:** Need to define the assistant's role, operational limits, safety rules, and appropriate use of messaging and patient data.

| ID | Code | Operational definition | Illustrative excerpt (translated) |
|---|---|---|---|
| P14 | **Governance and scope of the assistant/chat** | Apply when the participant questions the role of the AI assistant or messaging function, or requests protocols, limits, caution, objectivity, safety, or rules for their use. | *"The assistant and direct chat require well-defined protocols to ensure the security of patient data."* |

**P-T4. Systemic integration as the basis for continuity of care**

**Theme definition:** Captures interoperability with existing health-information systems and the role of shared, traceable records in reducing duplicate work and supporting multiprofessional continuity.

**P4-A. Interoperability and reduction of duplicate work**

**Category definition:** Integration with PEC/e-SUS or other systems to avoid repeated entry, transcription, and fragmented screens.

| ID | Code | Operational definition | Illustrative excerpt (translated) |
|---|---|---|---|
| P18 | **Interoperability with PEC/e-SUS and reduction of duplicate work** | Apply when the participant recommends integration with PEC/e-SUS or other systems to avoid duplicate entry, multiple screens, manual transcription, or rework. | *"It would greatly help the multiprofessional team if these data populated the patient history without duplicate entry."* |

**P4-B. Multiprofessional continuity and coordination**

**Category definition:** Shared information and aligned goals across professionals and encounters.

| ID | Code | Operational definition | Illustrative excerpt (translated) |
|---|---|---|---|
| P19 | **Continuity and multiprofessional communication** | Apply when the participant reports that the platform or its integration could support care coordination, communication among professionals, aligned goals, or continuity of care. | *"It would reduce communication gaps and ensure that the goal I agreed with the patient is the same one the nutritionist reinforces later."* |

**P4-C. Information governance and official record**

**Category definition:** Security, traceability, and the legitimacy of formal records in shared care.

| ID | Code | Operational definition | Illustrative excerpt (translated) |
|---|---|---|---|
| P20 | **Information security and official record** | Apply when the participant emphasises official documentation, information security, traceability, formal recording of actions, or greater reliability of data in shared care. | *"The impact would be on information security."* |
