## Supplementary File S8. Item-level System Usability Scale (SUS) scores among users and health professionals for "Boora, an AI-assisted digital platform for overweight and obesity care in Brazilian primary care: a formative mixed-methods evaluation of perceived usability and acceptability" (from another version of this preprint: version 1, 2026-07-16)

| No. | System Usability Scale item | Wording | Users (n = 15) Adjusted score, mean (SD) | Health professionals (n = 8) Adjusted score, mean (SD) |
|---|---|---|---|---|
| 1 | I think that I would like to use this system frequently. | Positive | 3.27 (0.46) | 3.50 (0.53) |
| 2 | I found the system unnecessarily complex. | Negative | 3.13 (0.35) | 3.00 (0.00) |
| 3 | I thought the system was easy to use. | Positive | 3.27 (0.46) | 3.25 (0.46) |
| 4 | I think that I would need the support of a technical person to be able to use this system. | Negative | 2.67 (0.72) | 2.75 (0.46) |
| 5 | I found the various functions in this system were well integrated. | Positive | 3.13 (0.35) | 3.13 (0.35) |
| 6 | I thought there was too much inconsistency in this system. | Negative | 3.13 (0.35) | 3.00 (0.00) |
| 7 | I would imagine that most people would learn to use this system very quickly. | Positive | 3.13 (0.64) | 3.50 (0.53) |
| 8 | I found the system very cumbersome to use. | Negative | 3.13 (0.35) | 3.00 (0.00) |
| 9 | I felt very confident using the system. | Positive | 2.87 (0.52) | 3.00 (0.00) |
| 10 | I needed to learn a lot of things before I could get going with this system. | Negative | 2.87 (0.64) | 2.88 (0.35) |

**Notes.** SD, standard deviation. The SUS was administered in a cross-culturally adapted Brazilian Portuguese version (reference 21 in the main manuscript); items are shown here in the original English wording for reference.

Each item was answered on a five-point scale (1 = strongly disagree to 5 = strongly agree). Responses were then adjusted to a 0 to 4 scale on which a higher value always indicates better perceived usability: for positively worded (odd-numbered) items the adjusted score is the response minus 1, and for negatively worded (even-numbered) items it is 5 minus the response. Summing the ten adjusted item means and multiplying by 2.5 reproduces the mean total SUS score for each cohort.

All participants answered all ten items; there were no missing data. Standard deviations of 0.00 among health professionals reflect identical responses from all eight participants on those items.

Total SUS score, mean (SD): users 76.5 (10.3); health professionals 77.5 (4.6). Internal consistency (Cronbach alpha), computed on the adjusted item scores: users 0.94; health professionals 0.71.

In both cohorts the lowest adjusted item scores were for item 4 (needing the support of a technical person) and item 10 (amount to learn before use); among users, item 9 (confidence in use) was also comparatively low.

These item-level results are descriptive; no inferential testing was performed, given the small formative sample.
